# Absence of phosphorylated CDK4, a biomarker of intrinsic resistance to CDK4/6 inhibitors in head and neck squamous cell carcinoma, can be predicted by their gene expression profile

**DOI:** 10.1101/2025.11.10.25339594

**Authors:** Eric Raspé, Katia Coulonval, Jaime M. Pita, Alex Spinette, Ligia Craciun, Flavienne Sandras, Frederick Libert, Anne Lefort, Pieter Demetter, Gabrielle van Caloen, Likun Zhang, Wubin Quian, Sheng Guo, Denis Larsimont, Xavier Bisteau, Jean-Pascal Machiels, Pierre P. Roger

## Abstract

The level of T172 CDK4 phosphorylation, required for enzyme activity and entry in the normal cell cycle, was variable in two head and neck squamous carcinoma (HNSCC) cohorts, corresponding cell lines and patient-derived tumor xenograft (PDTX) models. Actively proliferating tumors or models lacking CDK4 phosphorylation had defective pRb, elevated *E2F1* or *CCNE1* expression or were HPV-positive. CDK4 phosphorylation absence was perfectly predicted with a gene expression-based tool developed to predict the CDK4 phosphorylation status in breast tumors after correction of two confounding factors (the *CDKN2A* mutation status and high *CDKN2A* locus expression with exclusive p14-coding mRNA expression). Phosphorylated CDK4 was detected in all cell lines or PDTX sensitive to CDK4/6 inhibitors while models lacking CDK4 phosphorylation were insensitive to them. Growth *in vivo* of a PDTX model and of the SCC9 cell line xenograft with phosphorylated CDK4 was not affected by treatment with these drugs. Lack of phosphorylated CDK4 informs thus on the irreversible intrinsic resistance to CDK4/6 inhibitors. Its prediction with a gene expression-based tool may guide inclusion of these drugs in HNSCC treatment.

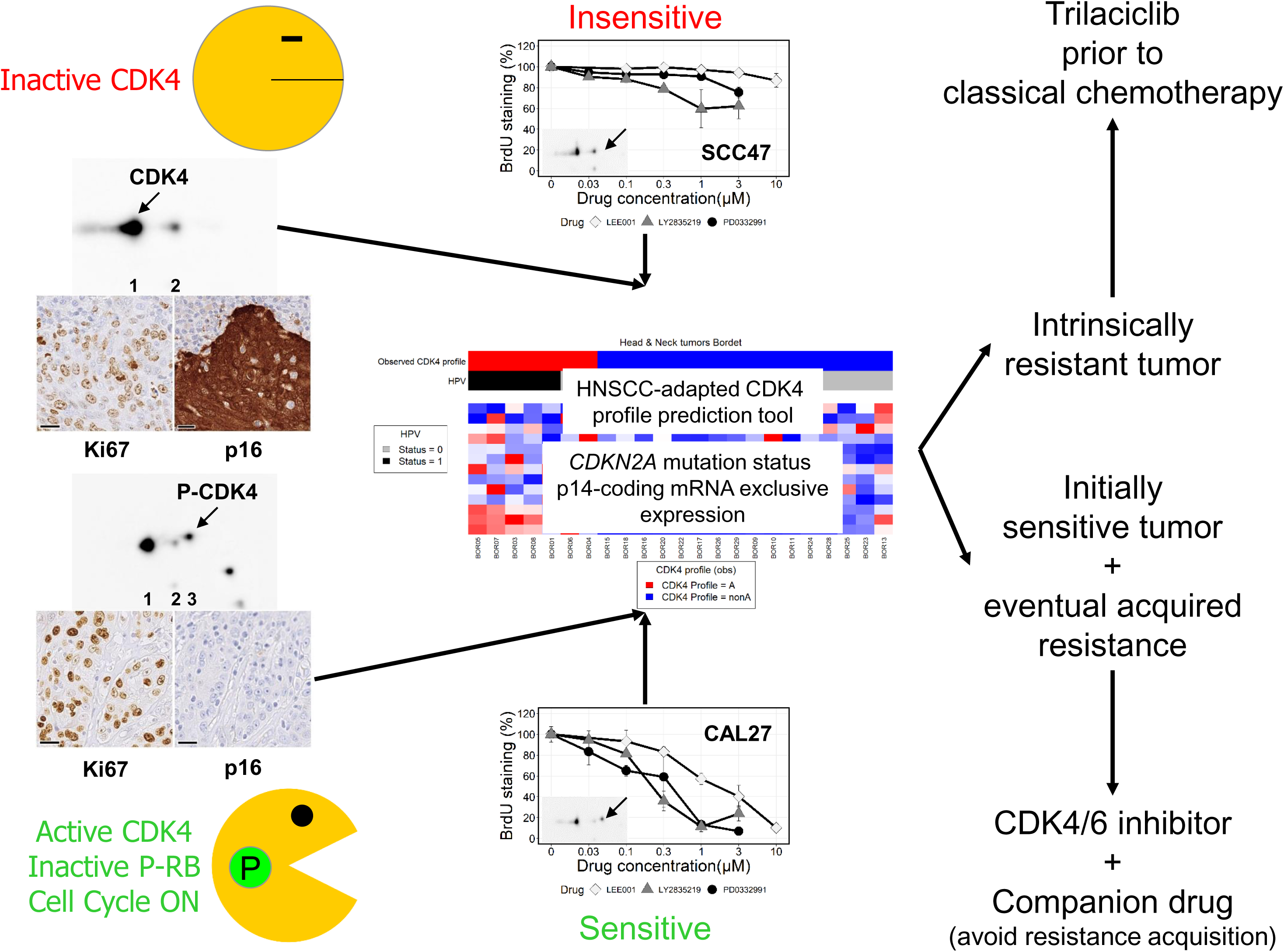

## Introduction

Head & neck squamous cell carcinoma (HNSCC) is the eight most common cancer worldwide with about 750 000 new cases and 360 000 fatalities each year (Sung, Ferlay et al., 2021). It originates from the epithelial lining of the oral cavity, oropharynx, larynx or hypopharynx and is generally due to tobacco and/or alcohol consumptions (Leemans, Snijders et al., 2018). High risk human papillomaviruses (HPVs) cause oropharyngeal cancer (OPC), a distinct disease with its own molecular profile and clinical characteristics including better prognosis and good response to combined radiotherapy and cisplatin-based chemotherapy (Lawrence, 2015, Leemans et al., 2018). Single surgery or radiotherapy is usually offered to HNSCC patients with early-stage disease with a cure rate ranging between 70 to 90%, whereas patients with locally advanced disease receive multimodal treatments that include surgery followed by (chemo)-radiation or primary chemo-radiation with a cure rate of about 50% (Machiels, René Leemans et al., 2020). As the cure rate of advanced disease is still low, there is a need to improve the treatment efficacy.

Cell cycle entry dysregulation is key to cancer (Hanahan & Weinberg, 2011). Mitogenic signalling leads to increased expression of D-type cyclins, which associate to CDK4 and CDK6 kinases triggering their activation. These formed complexes in turn phosphorylate and thereby inactivate the pRb tumor suppressor (Malumbres & Barbacid, 2009, Sherr & McCormick, 2002). When unphosphorylated, pRb sequesters E2F1/2/3 transcription factors, blocking the expression of key genes involved in the control or execution of the cell cycle. Upon commitment to cycle, pRb phosphorylation is maintained by a positive feedback loop linking pRb to E2F-dependent transcription of cyclin E1, which activates CDK2, and further phosphorylates pRb. The CDK4 activity is diversely regulated. It requires the phosphorylation of the enzyme on T172 (Kato, Matsuoka et al., 1994) but can also be blocked by CDK4 inhibitors like p16 (*CDKN2A-D* genes), which interferes with the binding of D-type cyclins to this kinase (Asghar, Witkiewicz et al., 2015). Our group has identified this cyclin D-bound CDK4 activating T172-phosphorylation as the last distinctly regulated step in CDK4 activation, determining the cell cycle commitment in pRb-proficient cells (Bockstaele, Bisteau et al., 2009, Bockstaele, Coulonval et al., 2006a, Bockstaele, Kooken et al., 2006b, Colleoni, Paternot et al., 2017, Paternot, Bockstaele et al., 2010, Paternot & Roger, 2009). In breast cancer tumors and cell line models, the phosphorylated CDK4 abundance is highly variable. Its presence or absence is associated respectively with the sensitivity or insensitivity of breast or thyroid cancer and mesothelioma cell lines to CDK4/6 inhibitors like palbociclib (Paternot, Raspé et al., 2024, Pita, Raspé et al., 2023, Raspe, Coulonval et al., 2017).

The HNSCC molecular landscape (Leemans et al., 2018) specifically affects the control of their cell cycle as the *RB1* tumor suppressor gene is often intact in HPV-negative tumors that also generally lack *CCNE1* or *CCNE2* gene amplification. In HPV-negative disease, alterations (mutations or gene copy number variations) are often detected upstream of *RB1* and mostly common in the *TP53* (84%), *CDKN2A* (58%), *PIK3CA* (34%) and *CCND1* (31%) genes. In HPV-infected tumors, however, E6 and E7 viral proteins triggers the degradation of p53 and pRb proteins, respectively, leading to uncontrolled cell cycle entry and increased survival of the infected cells (Graham, 2017). Thus, the molecular landscape of HNSCC, compatible with the activity of CDK4/6 inhibitors in HPV-negative HNSCC, provides a strong rationale to evaluate whether their efficacy to treat advanced ER-positive breast cancer (Goel, Bergholz et al., 2022) could be extended to these tumors. Several recent encouraging preclinical studies have confirmed that such inhibitors block the cell cycle entry in HNSCC cell lines *in vitro* (Beck, Georgopoulos et al., 2016, Gadsden, Fulcher et al., 2021, Hu, Peng et al., 2020, Ku, Yi et al., 2016, van Caloen, Schmitz et al., 2020, van Caloen, Schmitz et al., 2021, Zainal, Lee et al., 2019) or delay HNSCC patient-derived tumor xenograft (PDTX) model growth *in vivo* (Karamboulas, Bruce et al., 2018, van Caloen et al., 2020, van Caloen et al., 2021). Yet, the Palatinus (Adkins, Lin et al., 2021) and the NCT021034 (Oppelt, Ley et al., 2021) trials failed to demonstrate a statistically significant benefit of treating patients with the combination palbociclib/cetuximab versus the use of cetuximab alone in patient with HPV-negative progressive disease after first line therapy. Abemaciclib appeared also unbeneficial on a subset of platinum-refractory HNSCC (Keam, Hong et al., 2024). However, a promising objective response rate was recently reported upon palbociclib treatment of a small cohort of naïve untreated HPV-negative HNSCC, particularly if they had *CDKN2A* alterations (Oppelt, Ley et al., 2025). A better understanding of the intrinsic or acquired resistance to these drugs in HNSCC is thus required to either better select patients who will benefit from including CDK4/6 inhibitors in their treatment or to adapt it by appropriately combining these inhibitors with other drugs.

The aim of this work was to determine if the CDK4 T172-phosphorylation level is also variable in HNSCC and could inform on their sensitivity or resistance to CDK4/6 inhibitors. As CDK4 T172-phosphorylation status in breast tumors can be predicted by a surrogate tool using their gene expression profile, we also verified if this tool can be applied on HNSCC to identify patients likely to benefit from inclusion of CDK4/6 inhibitors in their treatment. If confounding factors (*CDKN2A* mutations or exclusive high expression of the *CDKN2A* mRNA coding for the p14 protein) affecting this prediction are considered, the tool perfectly predicted whether or not CDK4 is phosphorylated. As preclinical models without phosphorylated CDK4 are insensitive to CDK4/6 inhibitors, this tool is likely to identify patients who may benefit from inclusion of CDK4/6 inhibitors in their treatment.

## Results

### CDK4 phosphorylation is variable in HNSCC tumors

Proteins extracted from flash-frozen samples of HNSCC were analyzed by 2D gel electrophoresis and Western Blotting using an antibody specific to CDK4 to detect its modifications and evaluate the variations of the CDK4 phosphorylation level in these tumors. Typical CDK4 modification profiles, illustrated in Figure 1, revealed three main forms of CDK4 in HNSCC tumors similar to those observed in other cell models (Bisteau, Paternot et al., 2013, Bockstaele et al., 2006b, Raspe et al., 2017). The first most basic form is the native unmodified CDK4. The second form is neither labeled by ^32^P nor recognized by anti-T172- phosphorylated CDK4 antibodies in contrast to the third most acidic form of CDK4 indicated by an arrow on the right of the profiles which corresponds to T172-phosphorylated CDK4 (Bockstaele et al., 2006b, Coulonval, Vercruysse et al., 2022). The level of CDK4 phosphorylation was variable in HNSCC and associated to specific p16/Ki67 IHC staining profiles as illustrated in Figure 1. The CDK4 phosphorylation was not observed in some tumors despite their active phosphorylation indicated by their numerous Ki67-positive cells. These tumors, often HPV-positive, also displayed strong positive p16 IHC staining. By contrast, the CDK4 phosphorylation was detected and sometimes the most abundant modified form of the enzyme in HPV-negative, Ki67-positive, p16-negative tumors.

**Figure 1.**
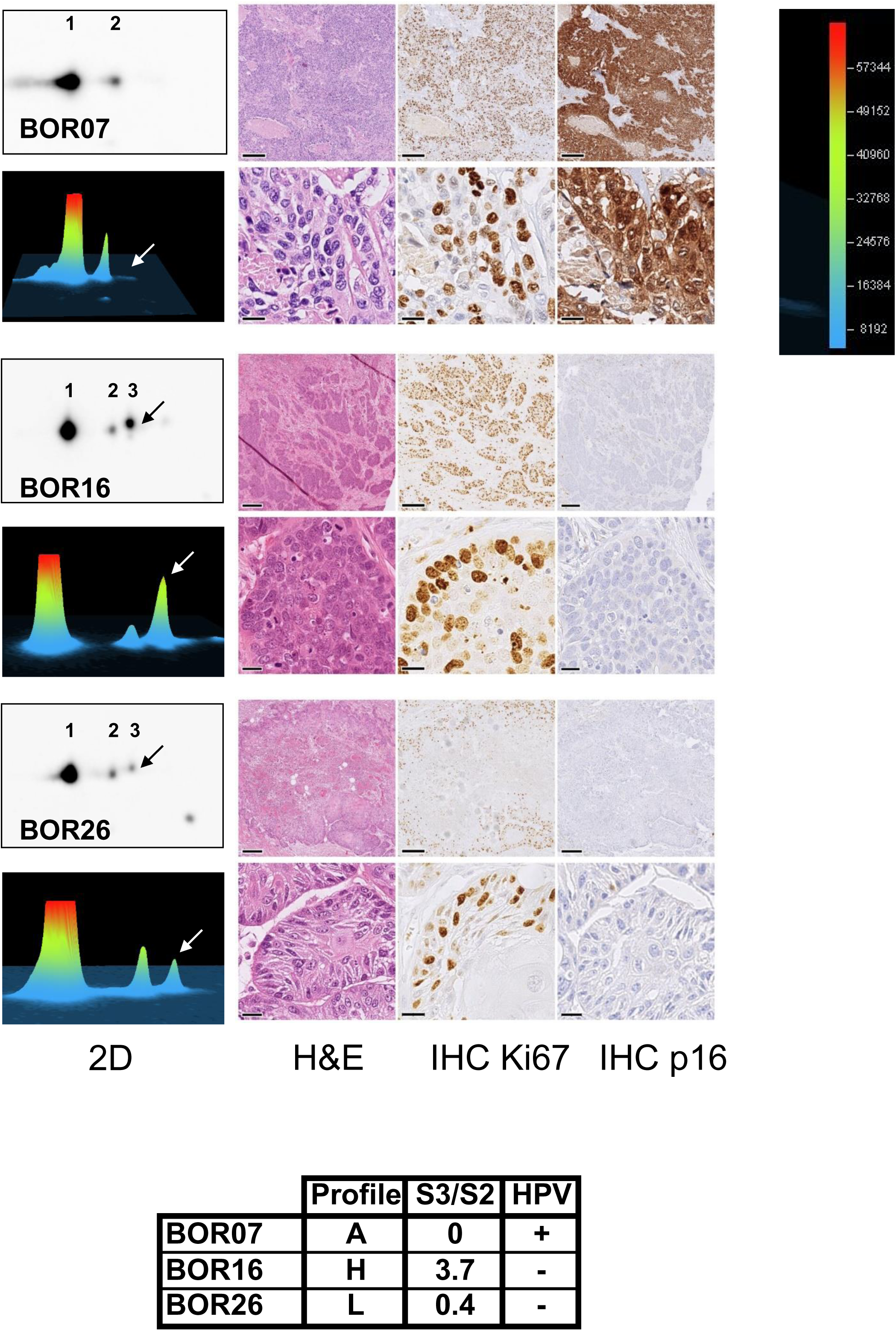
Characterization of the CDK4 modification profiles of human HNSCC. Total proteins extracted from the indicated tumors were separated by 2D electrophoresis and revealed by Western blotting with an anti-CDK4 antibody. Chemiluminescence images of the Western blots acquired with a Vilber-Lourmat Solo7S camera are shown on the left of the figure. These images were scaled to get a constant distance measured with the Qpath software between spot1 (native CDK4) and the first modified form of CDK4 (spot2). A 300 x 200 pixels image centered on the spot2 center of mass coordinates was extracted from the whole scaled image with the magick library in R. The phosphorylated CDK4 signal (spot 3) is indicated by an arrow. The CDK4 spot volume quantifications with the Bio1D software are shown below with the intensity scale on the right of the figure. The spot3/spot2 (S3/S2) ratio values, the HPV status and the types of CDK4 profile are indicated in the lower table. A single protein preparation was used for each tumor. Representative regions of interest of HE staining, Ki67 IHC staining and p16 IHC staining, respectively, of consecutive sections of the indicated tumors are displayed aside the 2D electrophoresis profiles. The scale bars on the lower left corner of each image of the upper row indicates 200 µm. The ones of the lower row indicates 20 µm. A single Ki67 or p16 IHC and a single HE staining were prepared for each tumor.

### Phosphorylated CDK4 in HNSCC cell lines, its relations to cell cycle regulator expression and CDK-containing complexes pRb-kinase activity, and sensitivity to the drugs

To correlate the variations of the CDK4 phosphorylation with the sensitivity to CDK4/6 inhibitors, proteins were extracted from our initial selection of untreated growing HNSCC cell lines (van Caloen et al., 2020) and analyzed by 2D gel electrophoresis and Western Blotting using an antibody specific to CDK4. The cell cycle entry rate was evaluated in parallel cell cultures with a 96-well plate BrdU incorporation assay. Figure 2A confirms that ribociclib (LEE011) dose-dependently reduced the proportion of cell labelled by a BrdU pulse in the HPV-negative cell lines (Cal27, Detroit 562, FaDu, SSC9, SCC15 and SCC25) but not in the HPV-positive cell lines SSC47 and SCC90 as did the two other CDK4/6 inhibitors, palbociclib (PD0332991) and abemaciclib (LY2835219). A rebound of BrdU incorporation was noticed at the highest abemaciclib concentration particularly in Detroit 562 cells and to a lesser extend in Cal27 and FaDu cells (Figure 2A) as well as in SCC9 cells treated with the highest concentration of palbociclib. As reduction of the MTT activity (Figure 2B) was observed in parallel in these cell lines, a cytotoxic activity presumably linked to higher DNA damage and repair labelled by the BrdU pulse may explain this paradoxical increase in BrdU labelling. The CDK4 modification profiles of the corresponding cell lines are displayed as large inserts in Figure 2A and as small flipped inserts on top of Figure 3A. Phosphorylated CDK4 was detected in all cell lines sensitive to CDK4/6 inhibitors but not in the two HPV-positive cell lines insensitive to the drugs. The expression of key cell cycle proteins was evaluated in total protein extracts of the 8 studied HNSCC cell lines (Figure 3A) by Western blotting. CDK4, cyclin D3 and cyclin A2 were expressed in all cell lines with limited variations in contrast to the more variable expression of CDK6, Cyclin D1, Cyclin E1, pRb, p21, p53 and of the p14 and p16 proteins. CDK6 was expressed in all but the SCC90 cells with the highest level in Cal27 and Detroit 562 cells. Cyclin D1 was also expressed in all models but at more variable levels. pRb and phosphorylated pRb were expressed roughly at the same level in all cells except the two HPV-positive ones due to the degradation of the pRb protein mediated by the E7 viral protein, as expected. The endogenous CDK4/6 inhibitor p16 and the protein p14 (encoded by the *CDKN2A* locus on an alternative reading frame by an alternative transcript) were only strongly expressed in the two HPV-positive models. The pRb kinase activity and composition of protein complexes isolated in non-denaturating conditions and immunoprecipitated with antibodies specific to cyclin D1, p21, CDK4, phosphorylated CDK4 (pCDK4) and CDK6 was evaluated *in vitro* by incubating the purified complex samples with a pRb fragment and ATP (Figure 3B). A variable pRb kinase activity was precipitated with the anti-cyclin D1, anti-CDK4, anti-CDK6 and anti-p21 antibodies from the different cell line extracts. Extracts from the HPV-positive SCC47 and SCC90 cell lines in which no or very little CDK4 phosphorylation was detected and in which the CDK4 was associated with the p16 protein, but not with cyclin D1, had no pRb-kinase activity. The SCC9 cell extracts purified with the anti-cyclin D1, anti-CDK4 and anti-CDK6 antibodies were much less active than the other active extracts and lacked both p21 and cyclin D1. This cell line which continued to grow *in vivo* despite treatment of the mice with ribociclib (van Caloen et al., 2020) and showed the largest residual proliferation rate *in vitro* upon exposure to CDK4/6 inhibitors, displayed the highest level of cyclin E1 (Figure 3A). The pRb kinase activity of CDK4 complexes was also proportional to the level of phosphorylated CDK4 after purification with anti-cyclin D1, anti-p21 and anti-CDK4 antibodies, except in the FaDu cells. In this cell line, a higher presence of CDK4 phosphorylation compared to pRb kinase activity was likely explained by CDK4 binding to p27 instead of p21 (Figure 3B). Indeed, we previously observed that p27 binding may inhibit the activity of CDK4 while stabilizing its T172 phosphorylation (Bockstaele et al., 2006b, Colleoni et al., 2017, Coulonval et al., 2022). Overall, this survey revealed that in CDK4/6 inhibitor-sensitive cells the pRb-kinase activity of CDK4 and CDK6 is associated to their binding to cyclin D1 and p21 as well as CDK4 phosphorylation. By contrast, in HPV-positive cells, the insensitivity to CDK4/6 inhibitors is not only associated with a low pRb presence, but also to the inactivity of CDK4 and CDK6 that are bound to p16 instead of cyclin D1, which precludes the activating phosphorylation of CDK4.

**Figure 2.**
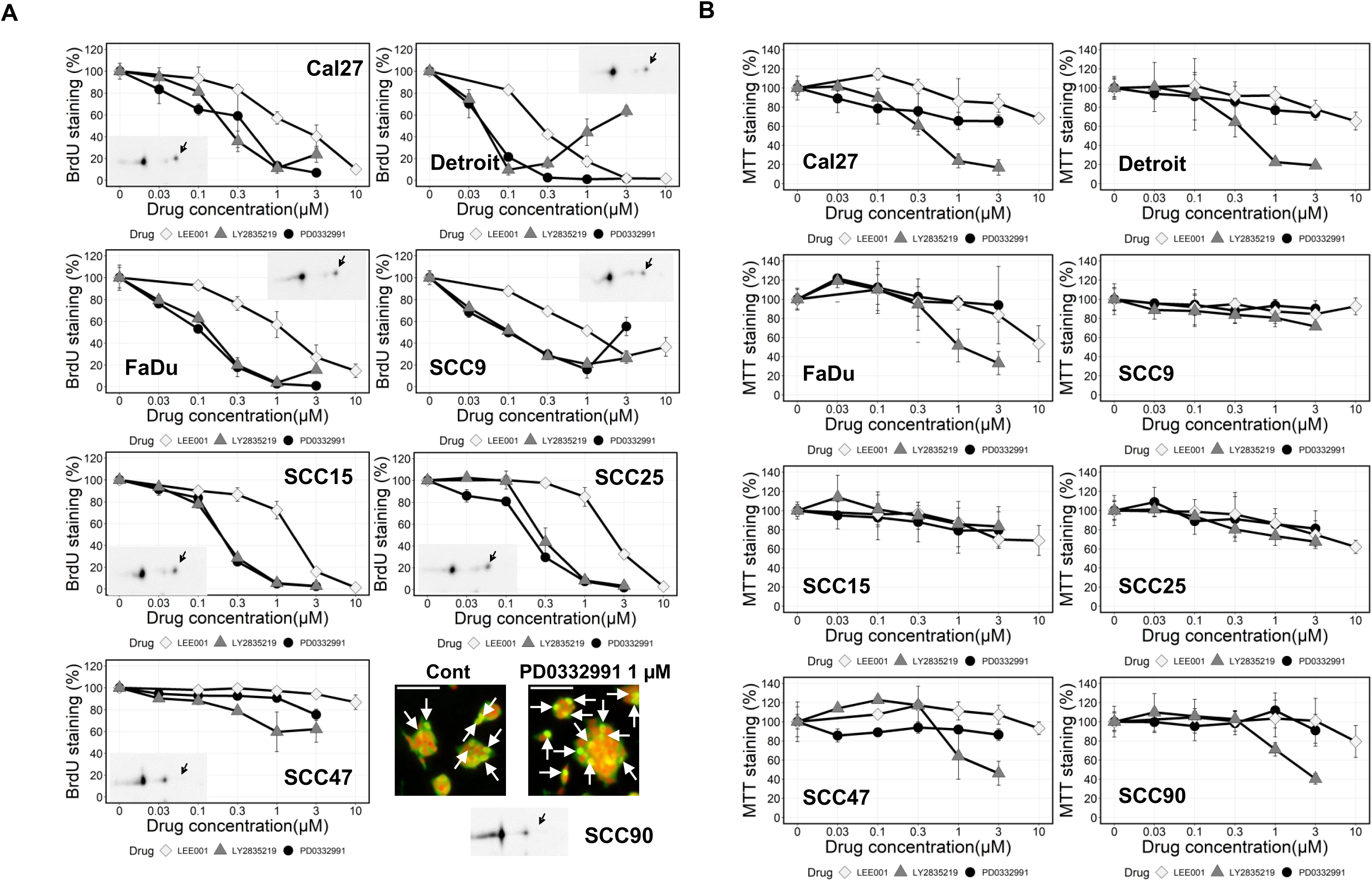
HNSCC cell line model characterization. A. The effects of increasing concentrations of ribociclib (LEE011, white diamonds), abemaciclib (LY2835219, grey triangles) or palbociclib (PD0332991, black circles) on the S-phase entry rate in the indicated cell line (Cal27, Detroit 562 (noted Detroit), FaDu, SCC9, SCC15, SCC25 and SCC47) was measured by counting the proportion of cell nuclei labeled by a BrdU pulse. Each drug concentration was tested in triplicate wells of a 96-well plate. The proportions of BrdU-labeled nuclei in a well were estimated in 10 fields per well and averaged. For each cell line and condition, the mean proportion of BrdU-labeled nuclei in these fields were averaged between the triplicate wells and displayed as means ± SD. The values presented are from a typical experiment done at the same date with the three drugs for each cell line. The number of experiment done with independent cultures of each cell line are reported in the Dataset EV3. The corresponding cell line CDK4 modification profile determined by 2D gel electrophoresis of total protein extracts revealed by Western blotting with an anti-CDK4 antibody are displayed as insert in the dose response graphs or below the images (SCC90). The phosphorylated CDK4 form is indicated by an arrow. As the UM-SCC090 cells (noted SCC90) grow as 3D clumps, staining image focalization was insufficient for the BrdU labeling quantification using the ImageJ script developed to this end. Instead, representative staining images are displayed. BrdU-labeled nuclei are indicated by the white arrows. The scale bars in the top left corners of the images indicates 100 µm. The UM-SCC090 CDK4 modification profile determined by 2D gel electrophoresis of total protein extract revealed by Western blot with an anti-CDK4 antibody is displayed below the images. A single protein extract was analyzed with a single 2D gel electrophoresis for each cell line. B.The impact of increasing concentrations of the indicated drugs on the mitochondrial activity measured by the MTT cell viability assay in the 8 tested cell lines are illustrated. Drugs codes are the same as in panel A. Data are means ± SD of the MTT activity relative to the control conditions computed with data from 3 independent experiments.

**Figure 3.**
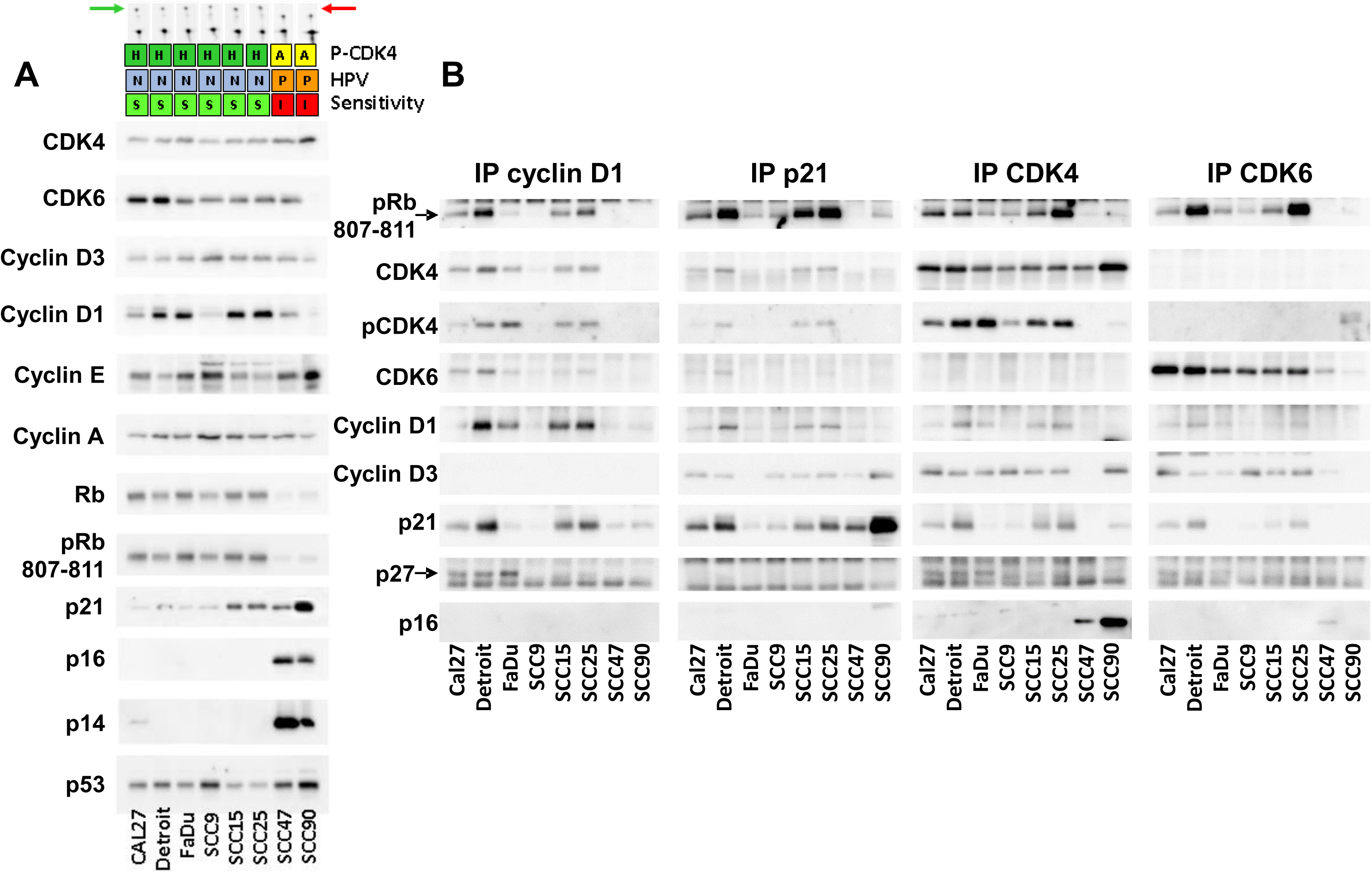
Characterization of the CDK complexes extracted from the HNSCC cell lines. A.The protein expression revealed by Western blotting of the total protein extract from the indicated cell lines with the indicated antibodies are illustrated. The two last cell lines are the HPV-positive models insensitive to CDK4/6 inhibitors. The phosphorylated CDK4 status, the HPV infection status and the *in vitro* sensitivity of the cell lines are color-coded on top of the Western blots. Smaller flipped images of the CDK4 2D profiles are illustrated on top of the figure. The green arrow points to the phosphorylated CDK4. The red arrow points to its absence. One gel per protein analyzed was loaded with the same protein extract for each cell line. B. Co-immunoprecipitation assays (IP) with an anti-cyclin D1, anti-p21, anti-CDK4 or anti-CDK6 antibody followed by *in vitro* pRb-kinase assay and western blotting immunodetection of the indicated proteins (CDK4; P-CDK4, T172-phosphorylated CDK4; CDK6; cyclin D1; cyclin D3). p27 is pointed by arrows in the corresponding immunoblots. One gel per protein analyzed was loaded with the same co-immunoprecipitated protein extract for each cell line.

### Phosphorylation of CDK4 in HNSCC is associated with their molecular features and a specific gene expression profile

We next compared the variations of the CDK4 phosphorylation level in HNSCC to parallel Ki67 and p16 IHC staining and tumor gene expression profiles quantified by RNASeq in samples from the two independent cohorts of tumors collected at the IJB and the UCLouvain to correlate these variations with the tumor molecular features. The more diverse cohort (IJB) was used as an exploratory cohort while the UCLouvain cohort was used to confirm and extend the observations. Individual CDK4 profiles of tumors from the IJB or UCLouvain cohorts, are illustrated in Figure EV1 and EV2, respectively, together with corresponding Ki67 and p16 IHC aligned fields from consecutive parallel FFPE tissue section from the IJB cohort. The expression of key genes involved in the control or execution of the cell cycle quantified by RNA-Seq is documented in Dataset EV11 and summarized as heatmaps (Figure 4 and Appendix Figure S2).

**Figure 4.**
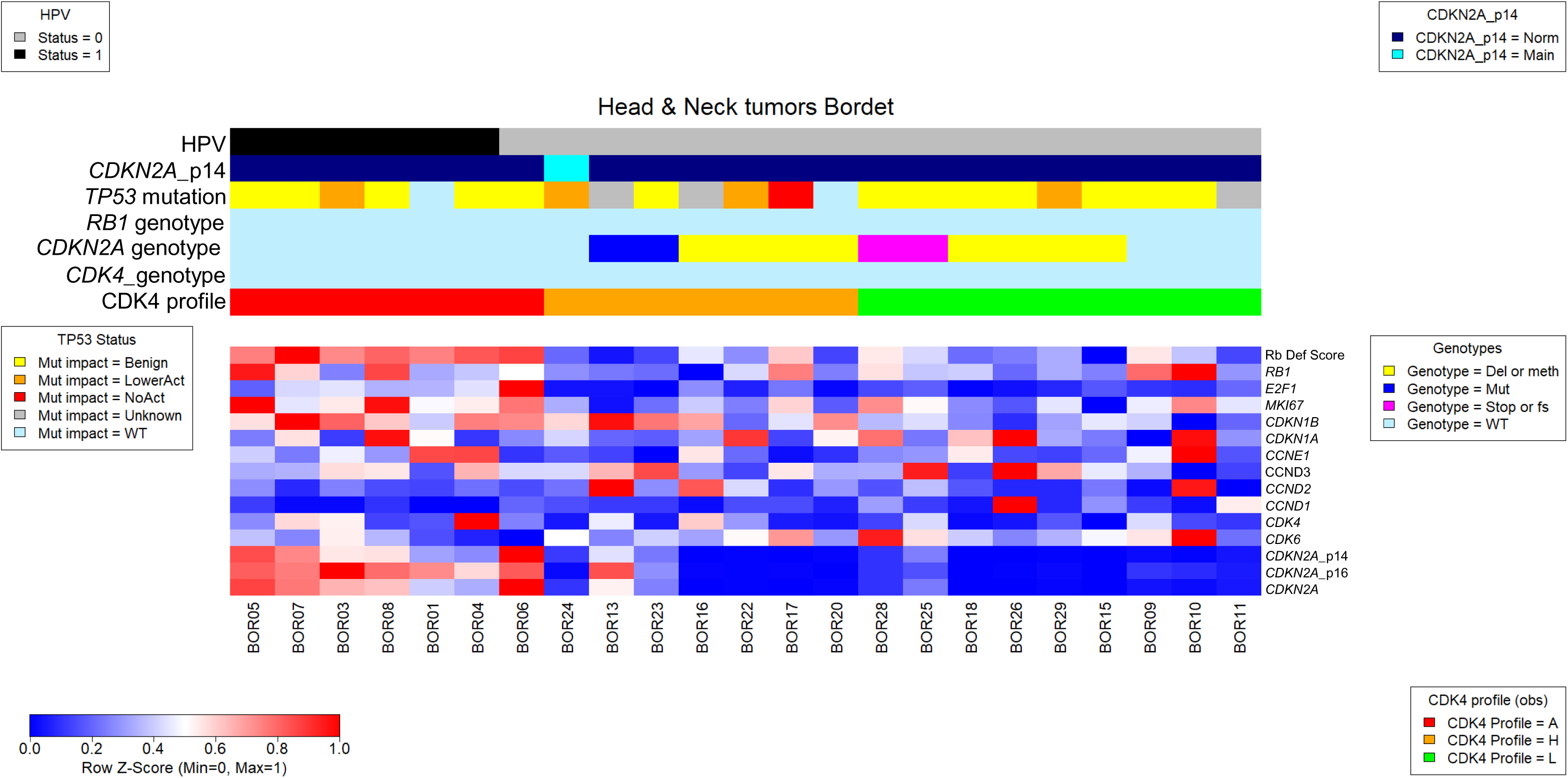
Expression of genes involved in the control or execution of the cell cycle in the human HNSCC of the IJB exploratory cohort. RNA was extracted in parallel from the same flash frozen tumor samples as those used to extract protein and quantified by RNA sequencing. The relative normalized expression values of the genes with the lowest value set to zero and the maximal value set to 1 are displayed as a heatmap drawn with the heatmap.plus R package as scaled in the bottom left panel of the figure. The names of the genes are indicated at the right of the heatmap. A single library was quantified for each tumor. The Rb deficiency index was computed as described previously (Raspe et al., 2017). For the *CDKN2A* gene, the global expression of the locus (noted CDKN2A) or the expression of the locus corrected by the contributions of the mRNA coding for p16 or p14 (noted *CDKN2A*-p14 and *CDKN2A*-p16, respectively) are displayed. The HPV infection (noted HPV) status, the p14 exclusivity (noted p14), the observed *TP53*, *RB1*, *CDKN2A* and *CDK4* genotypes and the observed CDK4 profile are color-coded on top of the heatmap with the features defined at the left of the color codes. The impact of the TP53 mutations is color-coded as defined in the legend on the left (NoAct = no activity due to Stop or frame shift mutations).

Despite very active proliferation suggested by positive Ki67 IHC staining, the CDK4 phosphorylation was absent in 7 tumors from the IJB cohort and in two tumors from the UCLouvain one. All proliferative tumors without phosphorylated CDK4 had high *CDKN2A* gene expression (Figure 4 and Appendix Figure S2) and stained positive for the p16 protein by IHC. HPV infection was revealed by alignment of RNA-Seq sequence reads from the tumors on sequences of the HPV genome in 7 of these tumors (6 in the IJB cohort and one in the UCLouvain one). The two HPV-negative tumor lacking phosphorylated CDK4 either expressed much more strongly the *E2F1* gene than all the other tumors from the cohort (BOR06, Dataset EV1) or had the lowest *RB1* gene expression (UCL30), Dataset EV1). CDK4 phosphorylation was detected in 31 tumors (16 from the IJB cohort and 25 from the UCLouvain cohort), all HPV-negative and with active proliferation. The p16 protein was generally not detected by IHC except in 3 tumors from the IJB cohort (BOR09, BOR10, BOR11) with phosphorylated CDK4 and normal *CDKN2A* gene expression. Phosphorylated CDK4 was detected in several tumors from both cohorts with elevated *CDKN2A* (BOR13, BOR23, BOR25 and BOR28 from the IJB cohort (Figure EV1) and UCL10, UCL15, UCL16, UCL47, UCL49, and UCL50 from the UCL cohort (Figure EV2)). Inspection of the read sequences in these tumors revealed the presence of mutations leading to a shorter dysfunctional protein or lying in the p16/CDK4 interface probably leading to altered interaction with CDK4. The p16 protein was detected by IHC in the BOR13 tumor with the G23D mutation coded by the exon 1α of the *CDKN2A* locus. As this mutation is located in the exon unique to the mRNA coding for the p16 protein, the p14 protein coded by an alternative transcript encoded by the *CDKN2A* locus on another reading frame remains intact. The p16 protein was not detected by IHC in the BOR28 tumor with the W110ter mutation, in the tumor BOR25 with the frameshift S56fs mutation as expected due to the truncation of the protein, as well as in the BOR23 tumor with the H83Y mutation (Figure EV1). The other tumors from the IJB cohort wherein CDK4 phosphorylation was detected were generally negative for p16 IHC staining. We noticed with the IGV tool that six samples form the UCL cohort (UCL11, UCL26, UCL28, UCL32, UCL41, UCL45) and one tumor from the IJB cohort (BOR24) displayed almost no coverage at the genomic location corresponding to the exon 1α of the gene coding for the start of the p16 protein (Figure EV3). The coverage values at exon 1α and exon 1β of the *CDKN2A* locus were extracted with a custom script in R to compute the relative contribution to the expression of the locus of the mRNA coding for p16 and p14. These are respectively represented as the p14 and p16 lines of the Figure 4 and Appendix Figure S2 heatmaps. All six of the HNSCC from the UCLouvain cohort with elevated *CDKN2A* gene expression had a relative contribution of the exon 1α coding for the p16 protein to the *CDKN2A* expression locus below 10% (skyblue code in the p14 row on top of the heatmap). Expression of the *CDKN2A* gene was generally normal (between 100 and 300 reads/20 million) or more often low (below 100 reads/20 million) in the other HNSCC with phosphorylated CDK4.

### Variations of the CDK4 phosphorylation in HNSCC PDTX models in relation to their molecular features and in vivo sensitivity to CDK4/6 inhibitors

To correlate the molecular features and the CDK4 phosphorylation levels to the *in vivo* sensitivity of HNSCC to treatment with CDK4/6 inhibitors, the CDK4 phosphorylation variations were analyzed in our initial small exploratory cohort of 5 PDTX models (Dataset EV2, worksheet “Selection QC PDTX UCLouvain”) characterized previously (van Caloen et al., 2020, van Caloen et al., 2021). To extend the validation of the impact of molecular features on the CDK4 phosphorylation, we completed this collection with a validation set of 25 Crown Bioscience PDTX models (Dataset EV2, worksheet “Selection QC PDTX Crownbio”). The CDK4 profiles of the models are documented, together with the expression of the Ki67 and p16 proteins evaluated by IHC staining of FFPE parallel tissue samples, in Figure EV4 and Figure EV5. The expression of selected genes involved in the control or the execution of the cell cycle quantified by RNA-Seq are summarized as a heatmap (Appendix Figure S3).

CDK4 phosphorylation was undetected in 8 models with high *CDKN2A* gene expression and strong p16 protein expression (1 from the UCLouvain and 7 from the Crown Bioscience cohorts) despite their active proliferation. Four of them (including one from the UCLouvain collection) were HPV-positive while the others had either elevated *E2F1* expression due to gene amplification, elevated *CCNE1* gene expression or defective *RB1* gene. CDK4 phosphorylation was detected in 22 models (4 from the UCLouvain collection and 18 from the Crown Bioscience cohort) with mutated *CDKN2A* or with intact *CDKN2A* locus and low or moderate *CDKN2A* gene expression. CDK4 was also phosphorylated despite high *CDKN2A* gene expression and positive p16 staining in a model with mutated *RB1* (HN2590) combined with the D84Y p16 mutation (Dataset EV2, worksheet “PDTX Crownbio”) affecting its binding to CDK4 (Ruas & Peters, 1998). Ribociclib treatment failed to control the growth of the HPV-positive UCLHN3 model in which CDK4 phosphorylation was undetectable. Ribociclib reduced the growth of 3 models out the 4 wherein CDK4 phosphorylation was detected (van Caloen et al., 2020, van Caloen et al., 2021).

### Prediction of the CDK4 modification profile in HNSCC tumors from the exploratory cohort

We have developed a tool to predict the CDK4 modification profile of a breast tumor using its gene expression profile (Raspe et al., 2017). The tool compares, gene by gene, the expression values of 11 genes of a tumor to references computed as means for each gene of their expression values in a collection of prototype tumors with a given CDK4 profile determined experimentally by 2D gel electrophoresis. In contrast to the breast tumors, distinction between H and L profile HNSCC was not associated with the vital status of the HNSCC patients (χ^2^ test p-value=0.59) nor with different means of the Rb deficiency index or of the expression of genes involved in the control or execution of the cell cycle except the *CDKN1B* gene (Appendix Table S1). Thus a binary prediction whether the CDK4 is phosphorylated (nonA) or not (A) was considered to test if the tool predicting the breast tumor CDK4 profile is applicable to HNSCC. The gene expression profiles of the 11 genes used to predict the CDK4 phosphorylation status of breast tumors are illustrated as heatmaps in the Appendix Figure S4 for the exploratory IJB or the validation UCLouvain tumor cohorts and for the Crown Bioscience PDTX models. These profiles are annotated with the observed and predicted CDK4 phosphorylation status on top of the heatmaps. Prediction errors were mainly false A profiles predictions associated with tumors or models with mutated CDKN2A gene or with high CDKN2A gene expression exclusive to the mRNA coding for the p14 protein. Table 1 summarizes the accuracies of the predictions by non-parametric and parametric versions of the tool according to the data sources and references used with individual profile observations and predictions reported in Appendix Table S2. Predictions were fully accurate in the exploratory IJB cohort using breast references when the *CDKN2A* gene expression values were corrected for the contribution of the p16-encoding mRNA to the locus expression and if the *CDKN2A* mutated tumors were excluded from the analysis. Predictions were perfect in this cohort with HNSCC references but only with the parametric rules. The tool performances were next validated with the expression data of the UCLouvain tumor cohort, the UCLouvain PDTX and the Crown Bioscience PDTX. In all cohorts, the best accuracy was achieved with a binary prediction using the parametric rule (difference between the correlation to the A and non-A references above or equal to a threshold defined with the IJB cohort), with the *CDKN2A* expression values corrected for the contribution of the p16-coding mRNA to the expression of the locus when tumors with mutated *CDKN2A* locus were excluded (Table 1).

**Table 1.** Accuracies of the prediction of the CDK4 phosphorylation status in the different tumor cohorts. The proportions (%) of tumors with correct prediction by the described tools are indicated for the HNSCC tumor cohort gathered at the IJB (noted Explo IJB), the initial HNSCC tumor cohort gathered at the Saint-Luc University Hospital of the UCLouvain (noted Validation UCLouvain) and for the Crown Bioscience cohort of 25 PDTX models (noted Validation CrownBio). The expression values used for the *CDKN2A* gene are either the raw counts normalized to the library size per 20 million reads (CP20M) or these values corrected for the contribution of the p16-coding mRNA to the expression of the *CDKN2A* locus (CP20M_p16). Five prediction tools are compared: a tool predicting the 3 profiles by comparison to references specific for each of them considering all samples (All); the same tool applied on the tumors without *CDKN2A* mutation (Cor Mut); and 3 binary tools predicting whether a tumor has a A or nonA profile. Two rules were considered: a tumor has a profile A in the first rule if the correlation to the profile A reference is the highest (noted Sup) or a tumor has a A profile if the difference between the correlation coefficients of the comparison between the expression of the test genes to the profile A or non-A references is above a threshold defined by a ROC analysis (noted ROCdelta). When the correlation coefficient of the comparison of the unknown sample gene expression profile to the H, L or nonA reference profiles is the same as the corresponding coefficient of the comparison to the A profile reference, the CDK4 modification profile cannot be determined. These cases are considered as false predictions in the computing of the accuracies.

|  |  | Binary | Binary | Binary | Binary | Binary | Binary |
| --- | --- | --- | --- | --- | --- | --- | --- |
|  |  | Breast | Breast | H&N | H&N | Breast | H&N |
|  |  | SupAHL | SupAHL | SupAHL | SupAHL | ROCdelta | ROCdelta |
|  |  | All | Cor Mut | All | Cor Mut | Cor Mut | Cor Mut |
| Explo<br>IJB | CP20M | 82.6 | **94.7** | **87** | **94.7** | **100** | **100** |
|  | CP20M_p16 | **95.7** | **100** | 78.3 | **89.5** | **100** | **100** |
| Validation<br>UCL | CP20M | 63.0 | 65 | **88.9** | **90** | 80 | 80 |
|  | CP20M_p16 | **88.9** | **90** | **88.9** | **100** | **95** | **100** |
| Validation<br>CrownBio | CP20M | 56 | 77.8 | 64 | 83.3 | **88.9** | 83.3 |
|  | CP20M_p16 | 72 | **94.4** | 72 | **94.4** | **100** | **100** |

## Discussion

In HNSCC tumors, cell lines and PDTX models as prototypes of tumors in which the frequency of mutated *CDKN2A* locus is high (Lawrence, 2015), the phosphorylated CDK4 level is variable. Some tumors or models completely lacked phosphorylated CDK4 despite their high proliferation rate. Furthermore, all models wherein CDK4 was not phosphorylated were insensitive to CDK4/6 inhibitors *in vivo* or *in vitro*. Tumors, cell lines and PDTX models without phosphorylated CDK4 share similar molecular defects as those identified in breast or thyroid tumors or in mesothelioma (Paternot et al., 2024, Pita et al., 2023, Raspe et al., 2017). Importantly, these defects are not restricted to defective pRb due to mutation, decreased pRb expression due to reduced gene copy number or protein degradation in HPV infected tumours mediated by the E7 protein encoded by the viral genome. We indeed extend here our observation that this lack of CDK4 phosphorylation can also occur and lead to resistance to CDK4/6 inhibitors even in the presence of intact pRb, namely when *CCNE1* or *E2F1* are overexpressed or amplified in tumors with intact *CDKN2A* locus (Paternot et al., 2024, Pita et al., 2023, Raspe et al., 2017). By similarity with the phenotype of the HNSCC cell lines, tumors lacking phosphorylated CDK4 are likely to be intrinsically resistant to the CDK4/6 inhibitors not only because pRb the target of CDK4/6 is missing or constitutively inactivated, but also because CDK4, one of the drug target, is itself inactive due to the absence of its T-loop phosphorylation (Bockstaele et al., 2006b, Kato et al., 1994). This association between the lack of CDK4 phosphorylation despite high proliferation rate and intrinsic resistance to the action of CDK4/6 inhibitors is thus a general rule, which identifies it as the most direct CDK4/6 inhibitor intrinsic resistance biomarker.

Although tumors or models lacking phosphorylated CDK4 have defective or constitutively inactivated pRb, not all models with altered pRb lack CDK4 phosphorylation. We identified one HNSCC PDTX with phosphorylated CDK4 despite that pRb is inactivated by a mutation. This model combines a pRb defect with mutated *CDKN2A* leading to loss of p16 function. Similarly, we identified several breast and mesothelioma cell lines insensitive to CDK4/6 inhibitors in which defective pRb or its low expression is combined with alterations of *CDKN2A* (gene deletion or mutation) and wherein CDK4 is phosphorylated (Paternot et al., 2024, Raspe et al., 2017). The detection of CDK4 phosphorylation in tumors or models in which pRb defects or *CCNE1* or *E2F1* amplification are combined with alteration of *CDKN2A*, and its absence in models with only pRb, *CCNE1* or *E2F1* defects, reveals that *CDKN2A* plays a key role in the control of CDK4 phosphorylation. The *CDKN2A* locus drives the expression of two transcripts through the use of two promoters sharing the last two exons and translated on different reading frames which encode two different proteins. These include p16 (which binds to and inhibits CDK4 and CDK6) and p14 (which binds to and inhibits MDM2) (Gil & Peters, 2006). A role for p14 in the CDK4 phosphorylation control is unlikely first because the tumor BOR13 with phosphorylated CDK4 as major modified form of the enzyme had high *CDKN2A* locus expression, stained positively for the p16 protein in IHC but bore a G23D mutation affecting the p16 binding to CDK4 (Scaini, Rossi et al., 2009). As this mutation lies in the first exon unique to p16, it does not affect the p14 protein. Second, in both cohorts of HNSCC tumors and in the PDTX model collections, CDK4 phosphorylation was still detected when tumors had a high *CDKN2A* locus expression associated with the exclusive expression of the p14-coding mRNA. This genetic evidence, consistent with recent reports (Li, Jiang et al., 2022, Palafox, Monserrat et al., 2022) and observations that the objective response rate to palbociclib treatment of untreated advanced HNSCC is higher when the *CDKN2A* gene is altered (Oppelt et al., 2025) and lower when p16 protein is detected (Dennis, Sacco et al., 2022), support the key role of *CDKN2A* in the control of CDK4 phosphorylation and its inclusion within the CDK4/6 inhibitor intrinsic resistance biomarkers.

Extending the indication of the CDK4/6 inhibitors to HNSCC will require a biomarker to identify patients who will likely benefit from the treatment. Reliable detection of CDK4 phosphorylation requires good quality proteins that can only be extracted from flash-frozen tissue not available in common clinical practice. We have previously developed a tool based on the gene expression profile of breast tumors to predict their CDK4 modification profile (Raspe et al., 2017) and now adapted it to predict the presence or absence of CDK4 phosphorylation in HNSCC. In the absence of *CDKN2A* mutation, adjusting the *CDKN2A* gene expression used in the tool by the relative contribution of the mRNA coding for the p16 protein perfectly corrected the CDK4 profile prediction of these HNSCC and their models. As similar high accuracies were achieved in two independent HNSCC cohorts and in the Crown Bioscience HNSCC PDTX cohort, the binary tool (prediction of presence or absence of CDK4 phosphorylation) is robust, independently validated and adapted to predict the HNSCC tumor and model intrinsic resistance to CDK4/6 inhibitors. On the other hand, because p16 staining was also observed in several tumors and models wherein CDK4 phosphorylation was detected including tumors or models with low or absent *CDKN2A* expression, p16 and Ki67 protein expression determined by immunohistochemistry alone may not predict robustly enough the CDK4 modification profile.

Despite their favorable genetic profile (low *RB1* gene alteration frequency, frequent *CCND1* gene amplification and defective *CDKN2A* gene), several trials failed to demonstrate a clear benefit of including CDK4/6 inhibitors in the treatment of HPV-negative HNSCC upon progression after cisplatin administration (Adkins et al., 2021, Keam et al., 2024, Oppelt et al., 2021). As reduced pRb expression was observed after cetuximab resistance acquisition (van Caloen et al., 2021) and because Robinson also observed *CCNE1* overexpression upon induction of cisplatin resistance in HNSCC models (Robinson, Rathore et al., 2019), the trial success may have been compromised because these defects were not tested beforehand to exclude likely unresponsive patients. Furthermore, cetuximab was probably not the appropriate drug to combine with palbociclib as its combination with ribocilib was less effective in controlling the HNSCC PDTX growth (van Caloen et al., 2020, van Caloen et al., 2021). Testing CDK4/6 inhibitors in patients with less advanced disease not exposed to platinum drugs may still be considered. Indeed, in a new trial testing palbociclib as single agent in untreated advanced HPV-negative HNSCC, the drug had a promising favorable impact, particularly in tumors with altered *CDKN2A* (Oppelt et al., 2025).

Detection or prediction of the CDK4 phosphorylation failed to predict the long-term *in vivo* response of one UCLouvain model treated with ribociclib. Treatment failure can be due to intrinsic resistance of the tumoral cells to the drug but also to a dynamic acquisition of drug resistance. As an example, even if the CDK4 is phosphorylated at baseline in SCC9 cells and if their entry in S phase is reduced *in vitro* by the three CDK4/6 inhibitors tested, ribociclib did not prevent their growth *in vivo* as xenografts in immuno-deficient mice (van Caloen et al., 2020). These cells kept a low residual S phase entry rate even at the highest drug concentrations tested. As these cells showed the highest expression of cyclin E1 of all HPV-negative cell lines and had low p21 expression, their CDK2 activity may bypass their CDK4/6 dependency and allow them to grow despite CDK4/6 inhibition. Exposure of Cal27 to palbociclib increased their *CCNE1* mRNA level and the cyclin E1 protein expression through the regulation of the eIF4I and eIF4G binding on the *CCNE1* mRNA, an effect blocked by the new mTOR inhibitor INK128 while overexpression of *CCNE1* gene and cyclin E1 protein in Cal27 and HN12 cells led to their resistance to palbociclib (Goto, Koshizuka et al., 2024). In other models as well, CDK2 and/or cyclin Es play a key role in the escape of the effects of CDK4/6 inhibitors (Asghar, Barr et al., 2017, Herrera-Abreu, Palafox et al., 2016, Knudsen, Kumarasamy et al., 2022, Raspe et al., 2017). Combined CDK4/6 and CDK2 inhibition improved the efficacy of the CDK4/6 inhibitor by preventing the acquisition of resistance to these drugs (Al-Qasem, Alves et al., 2022, Freeman-Cook, Hoffman et al., 2021, Goto et al., 2024, Pogacar, Johnson et al., 2022). We also observed that combining CDK4/6 inhibitors with MEK inhibitors improved the long term growth control of anaplastic thyroid cancer cell models (Pita et al., 2023). Combination with trametinib indeed prevented the upregulation of both Cyclins D and Cyclin E1, as well as the activation of ERK and mTOR pathways (as inferred by the decreased phosphorylation levels of ERK, p70S6K1, RPS6 and 4EB-P1) that increases the T172-phosphorylation of CDK4, which were observed upon palbociclib treatment. The HNSCC PDTX model able to grow upon treatment with CDK4/6 inhibitors despite its CDK4 phosphorylation described in our previous work may be a valuable resource to understand the mechanisms whereby HNSCC acquire resistance to CDK4/6 inhibitors and to test the above-mentioned drug combinations.

An alternative use of CDK4/6 inhibitors was recently approved by the FDA to treat small cell lung carcinomas (Dhillon, 2021). In this strategy, trilaciclib, a short-lived injectable CDK4/6 inhibitor is administered before treatment by classical chemotherapy. Trilaciclib induces a transient, reversible cell cycle arrest in normal haematopoietic stem and progenitor cells of the bone marrow, protecting them from damages during chemotherapy and thereby reducing the incidence of chemotherapy-induced myelosuppression. As cancer cells are not affected by trilaciclib, they remain sensitive to chemotherapy. Furthermore, in metastatic triple negative breast cancer, administration of trilaciclib prior to chemotherapy (gemcitabine+carboplatin) enhanced chemotherapy efficacy (Tan, Wright et al., 2022). Chemotherapy-based treatment is the recommended option for HPV-positive HNSCC from the oropharynx (Machiels et al., 2020). As these tumors lack phosphorylated CDK4, it would be worth testing if the trilaciclib-based strategy also enhances the efficacy of their chemotherapy-based treatment and if this strategy also improves the management of other HNSCC lacking phosphorylated CDK4. As intrinsic resistance of the tumor cell to the CDK4/6 inhibitor is key to the success of this alternative strategy, by identifying such tumors, the tool described in this work will be instrumental to select patients to be included in clinical trials testing the benefits of this alternative use of CDK4/6 inhibitors.

In conclusion, the CDK4 phosphorylation level is variable in HNSCC. Its lack indicates intrinsic resistance to CDK4/6 inhibitors in HNSCC models. This lack can be robustly and reproducibly predicted with the tool developed to predict the breast tumor CDK4 modification profile, provided that it is adapted to the HNSCC and that the tumor *CDKN2A* mutation status is defined. We clarified the rules to predict intrinsic resistance to CDK4/6 inhibitors by elucidating key confounding factors that hide the central role of p16 in this resistance.

## Materials & methods

### Cell lines and culture

Human head & neck squamous cell carcinoma cell lines (Cal27, FaDu, Detroit 562, SCC9, SCC15, SCC25, SCC47 and SCC90) are documented in Dataset EV3. They were shared by UCLouvain and authenticated by STR profiling by this laboratory (van Caloen et al., 2020). Cells were maintained in a humidified atmosphere (5% CO_2_) at 37°C and cultured as described previously (van Caloen et al., 2020). They were passaged for fewer than 2 months or 20 passages. Cell culture reagents were obtained from Gibco, (Carlsbad, CA, USA). Palbociclib (PD0332991; S1116), abemaciclib (LY2835219; S7158) and ribociclib (LEE011; S7440) were purchased from SelleckChem (Houston, TX, USA) and dissolved in DMSO. Controls were treated with the same concentration of DMSO.

### Patient-derived tumor xenograft models

The initial cohort of five PDTX models with characteristics summarized in Dataset EV2 used at the UCLouvain was described previously (van Caloen et al., 2020, van Caloen et al., 2021). Archival frozen tumor samples from untreated animals collected at the end of these experiments were used and retrospectively analyzed in this work. One sample per model was used except for model UCL02 for which a second independent sample was required due to a too high proportion of murine tissue in the initial sample revealed by the different migration profile of murine CDK4. Animal work was undertaken in compliance with Belgian laws and all experiments were in accordance with the UCLouvain local ethics committee (approval number: 2015/13AOU/445). Animal welfare is regularly controlled by inspections under Belgian laws, and all investigators performing animal work had successfully completed the Federation of European Laboratory Animal Science Associations (FELASA) Function C training. The protein extraction and 2D analysis were performed at the ULB with samples annotated only with the model identification number without knowledge of the model molecular profile. The RNA extractions were performed independently at the UCLouvain with samples annotated only with the model identification number without knowledge of the model molecular profile. RNA samples were provided to the ULB-BRIGHTCORE facility using a specific sample code for library preparation and sequencing. Alignment of the RNASeq sequences was also done at the ULB-BRIGHTCORE facility blinded with respect to the model molecular profiles. A validation cohort of flash-frozen PDTX samples collected in untreated animals was prepared from 25 models identified in the PDTX collection of Crown Bioscience (San Diego, CA, USA) and retrospectively analyzed in this work. Models were selected based on the data available in the Crown Bioscience database (https://hubase.crownbio.com/PDXmodel/HuPrime) to cover molecular profiles leading to the presence of CDK4 phosphorylation or its lack and eventual exceptions. The selection included one such model combining antagonistic alterations (*RB1* and *CDKN2A* gene mutations) likely leading to independence of CDK4 to enter the cell cycle together with an expected paradoxical detection of CDK4 phosphorylation. The selection further included 7 models wherein CDK4 phosphorylation is unexpected (3 HPV-positive models and 4 models with defects leading to a CDK4-independent cell cycle entry such as *CCNE1* or *E2F1* gene amplification or *RB1* gene deletion or mutation). The selection finally included 17 models wherein CDK4 phosphorylation is expected (6 models with mutated *CDKN2A*, 7 models with *CDKN2A* gene deletion or very low expression, 2 models with high expression of cell cycle genes and 2 models wherein these genes are moderately expressed). The protein extraction and 2D analysis were performed with samples annotated only with the model identification number without knowledge of the model molecular profile. The protein 2D analysis was run in 3 initial batches according to the model identification number. When required by the insufficient quality of the 2D gels or to confirm the CDK4 profile, the analysis was repeated with protein from the initial extraction. Alignment of the RNASeq sequences was also done blinded with respect to the model molecular profiles. The characteristics of the models and the model passage numbers are described in the Dataset EV2. All animal studies were approved by the Crown Bioscience Institutional Animal Care and Use Committee (IACUC) and conducted in their SPF facility in accordance with the NIH Guide for the Care and Use of Laboratory Animals. One frozen sample shipped on dry ice was analyzed per model. The numbers of protein or RNA extractions, 2D gel electrophoresis, hematoxylin/eosin, p16 or Ki67 staining per model are indicated in Dataset EV2.

### Selection and inclusion of patient samples

We retrospectively collected frozen HNSCC samples to investigate whether the CDK4 phosphorylation is detectable and variable in these tumors and to evaluate its clinical relevance. Parallel formalin-fixed, paraffin-embeded (FFPE) tissue was used for further characterization of the tumors by immuno-histochemistry (IHC). Based on our previous works on mesotheliomas, thyroid or breast tumors (Paternot et al., 2024, Pita et al., 2023, Raspe et al., 2017), at least 30 to 40 samples are required to find enough tumors wherein the CDK4 is not phosphorylated despite active proliferation. To gather an exploratory cohort as diverse as possible, we pre-selected from the Belgian register of biobanks, a first cohort of 40 HNSCC gathered between 2007 and 2020 including 7 HPV-positive cases in the collection of the Institut Jules Bordet (IJB) based on their reported HPV and p16 IHC status used to screen tumors for HPV infection (documented in the worksheet “Selection IJB” of Dataset EV4). After extraction of proteins and RNA from archival frozen samples of the selected tumors and their analysis by 2D gel electrophoresis and RNA-Seq, only 23 cases of the IJB cohort met the analyte quality criteria for both 2D gel detection of CDK4 and RNA extraction (as detailed in Appendix Supplementary Methods). The patient selection flow chart according to the specified criteria is described in Appendix Figure S1. The protein extraction and 2D analysis were performed at the ULB with samples annotated only with the tumor identification number without knowledge of the tumor molecular profile. The protein 2D analysis was run in 4 initial batches according to the tumor identification number. When required by the insufficient quality of the 2D gels or to confirm the CDK4 profile (as detailed in Appendix Supplementary Methods), the analysis was repeated either with protein from the initial extraction or from a new one. The RNA extractions were performed independently at the ULB by another scientist with samples annotated only with the tumor identification number without knowledge of the model molecular profile. RNA samples were provided to the ULB-BRIGHTCORE facility using a specific sample code for library preparation and sequencing. Alignment of the RNASeq sequences was also done at the ULB-BRIGHTCORE facility blinded with respect to the model molecular profiles. The initial sample characteristics together with the numbers of protein or RNA extraction, 2D gel analysis, RNASeq analysis or hematoxylin/eosin, p16 and Ki67 staining are documented in Dataset EV4 with their study inclusion status or cause of eventual rejection. The molecular features of the final tumor selection and informative clinical records of the corresponding patients are summarized in Dataset EV1 (worksheet “IJB cohort”). As the EHNS-ESMO-ESTRO Clinical Practice Guidelines do not recommend surgery for HPV-positive HNSCC, a second less diverse retrospective validation cohort was built with 72 archival HNSCC flash-frozen samples gathered between 2007 and 2019 obtained from the Cliniques universitaires Saint-Luc (UCLouvain cohort). The initial selection of the UCLouvain cohort was blinded with respect to the anatomical origin of the tumor. Six samples were later excluded for inappropriate anatomical location (2 tumors from the nasopharynx, 2 tumors from the external lower lip, 1 tumor from the skin and one lymph node metastasis of a tumor with the primary already included in the study). Only 27 cases from the UCLouvain cohort were similarly included in the study based on the quality criteria described above. The protein extraction and 2D analysis were performed at the ULB with samples annotated only with the tumor identification number without knowledge of the tumor molecular profile. The protein 2D analysis was run in 3 initial batches according to the tumor identification number. When required by the insufficient quality of the 2D gels or to confirm the CDK4 profile, the analysis was repeated either with protein from the initial extraction or from a new one. The RNA extractions were performed independently at the UCLouvain with samples annotated only with the tumor identification number without knowledge of the model molecular profile. RNA samples were provided to the ULB-BRIGHTCORE facility using a specific sample code for library preparation and sequencing. Alignment of the RNASeq sequences was also done at the ULB-BRIGHTCORE facility blinded with respect to the model molecular profiles. The initial sample characteristics together with the numbers of protein or RNA extractions, 2D gel analyses, RNASeq analyses or hematoxylin/eosin, p16 and Ki67 stainings are documented in the worksheet “Selection UCL” of the Dataset EV4 file with their study inclusion status or cause of eventual rejection. The patient selection flow chart is described in Appendix Figure S1. The tumor characteristics and the corresponding patient clinical records are summarized in Dataset EV1 (worksheet “UCLouvain cohort”). Collection of patient tissues and associated data was in agreement with the Ethics Committees of IJB (CE1978, CE2970) and UCLouvain (2018/13FEV/060) studied in accordance with the Declaration of Helsinki. As the study was retrospective and used residual human body material, asking informed consent of the patient was not required by the Ethics Committees. The demographic characteristics of the two final cohorts are compared in Appendix Table S3. Because HPV-positive tumors are generally treated by cisplatin-based chemotherapy coupled to radiotherapy at the UCL hospital while several HPV-positive tumors were collected at the IJB, the null hypothesis of dependence of the two cohorts cannot be rejected when relapse events, grade and binary CDK4 profile are considered due to their relation to the HPV status of the tumors. For the other clinical features, the two cohorts are similar.

### DNA synthesis and cell growth assay

The proportion of cells with active DNA synthesis and the effect of inhibitory drugs on cellular growth were respectively evaluated by the 5LbromoL2′Ldeoxyuridine (BrdU) incorporation assay and the sulforhodamine (SRB) and 3-[4,5-dimethylthiazol-2-yl]-2,5 diphenyl tetrazolium bromide (MTT) assays as described previously (Raspe et al., 2017) and in the Appendix Materials and methods.

### Protein analysis

Equal amounts of wholeLcell extract proteins or immunoprecipitates were separated by SDS-PAGE and immunodetected. For 2DLgel electrophoresis, cells were lysed in a buffer containing 7 M urea and 2 M thiourea while ground tissues obtained from cryogrinding of flash-frozen tumor tissues were solubilized and analyzed as described previously (Raspe et al., 2017) and in the Appendix Materials and methods. Co-immunoprecipitations were performed as described (Bockstaele et al., 2006b, Paternot, Coulonval et al., 2003) using the antibodies listed in Appendix Table S4 and the Structured Method reagent table. pRb-kinase activity of immunoprecipitated CDK complexes was measured by *in vitro* incubation with ATP and a fragment of pRb, as previously described (Paternot, Arsenijevic et al., 2006, Paternot et al., 2003).

### RNA extraction, sequencing and analysis

Total RNA from the IJB HNSCC ground tissues (obtained from cryogrinding of flash-frozen tumor pieces) and from OCT-embedded frozen tissue slides (at least, 10 sections of 10 μm per sample) were first extracted with TRI Reagent Solution (Invitrogen) using a Potter-Elvehjem homogenizer with a motorized PTFE pestle, and then purified with the RNeasy Mini kit (Qiagen) and on-column DNase digestion (RNase-free DNase Set, Qiagen) according to the manufacturer’s protocol. Total RNA were extracted from UCLouvain PDTX and tumors with phenol/chloroform and purification on RNeasy mini columns (Qiagen), according to manufacturer’s instructions. RNA yield and purity were assessed using a Fragment Analyzer 5200 (Agilent Technologies, Massy, France). Gene expression was quantified by RNASeq technology as described in detail in the Appendix Materials and methods. The Rb deficiency index was computed as described previously (Raspe et al., 2017). As detailed in the Appendix Materials and methods, the centroid method developed to predict the breast tumor CDK4 profile (Raspe et al., 2017) was applied to the HNSCC tumor gene expression quantifications to predict their CDK4 modification profile.

### Immunohistochemistry and tissue section analysis

Hematoxylin/eosin (HE) or KI67 and p16 immunohistochemical stainings of the IJB tumor cohort and of the UCLouvain PDTX models were performed at the Anatomopathology department of the IJB as described in detail in the Appendix Materials and methods. Analysis of sections of the UCLouvain tumor cohort were performed at the Anatomopathology department of the Saint Luc UCLouvain University hospital and scored by a pathologist. Analysis of FFPE tissue sections from Crown Bioscience head & neck PDTX model samples were performed using their routine procedure described in details in the Appendix Materials and methods.

### Data analysis, statistics presentation and availability

Statistical analyses were performed with R (version 4.2.3). To compare two independent groups, the non-parametric Wilcoxon test was used with the P<0.05 threshold used to reject the null hypothesis of equal means. Plots were drawn using the ggplot2 package in R. When the correlation coefficient of the comparison of the unknown sample gene expression profile to the H, L or nonA reference profiles is the same as the corresponding coefficient of the comparison to the A profile reference, the CDK4 modification profile cannot be determined. These cases are considered as false predictions in the computing of the accuracies. Characteristics and molecular data (for a limited selection of genes) of the PDTX models of the Crown Bioscience are available at the company website after registration. Extended molecular data can be acquired after approval by the company. Raw sequence data acquired with the RNA extracted from the tumors or PDTX of the UCLouvain and of the tumors and of the IJB were deposited at the EGA database with (submission ongoing).

## Structured Methods - Reagents and Tools Table

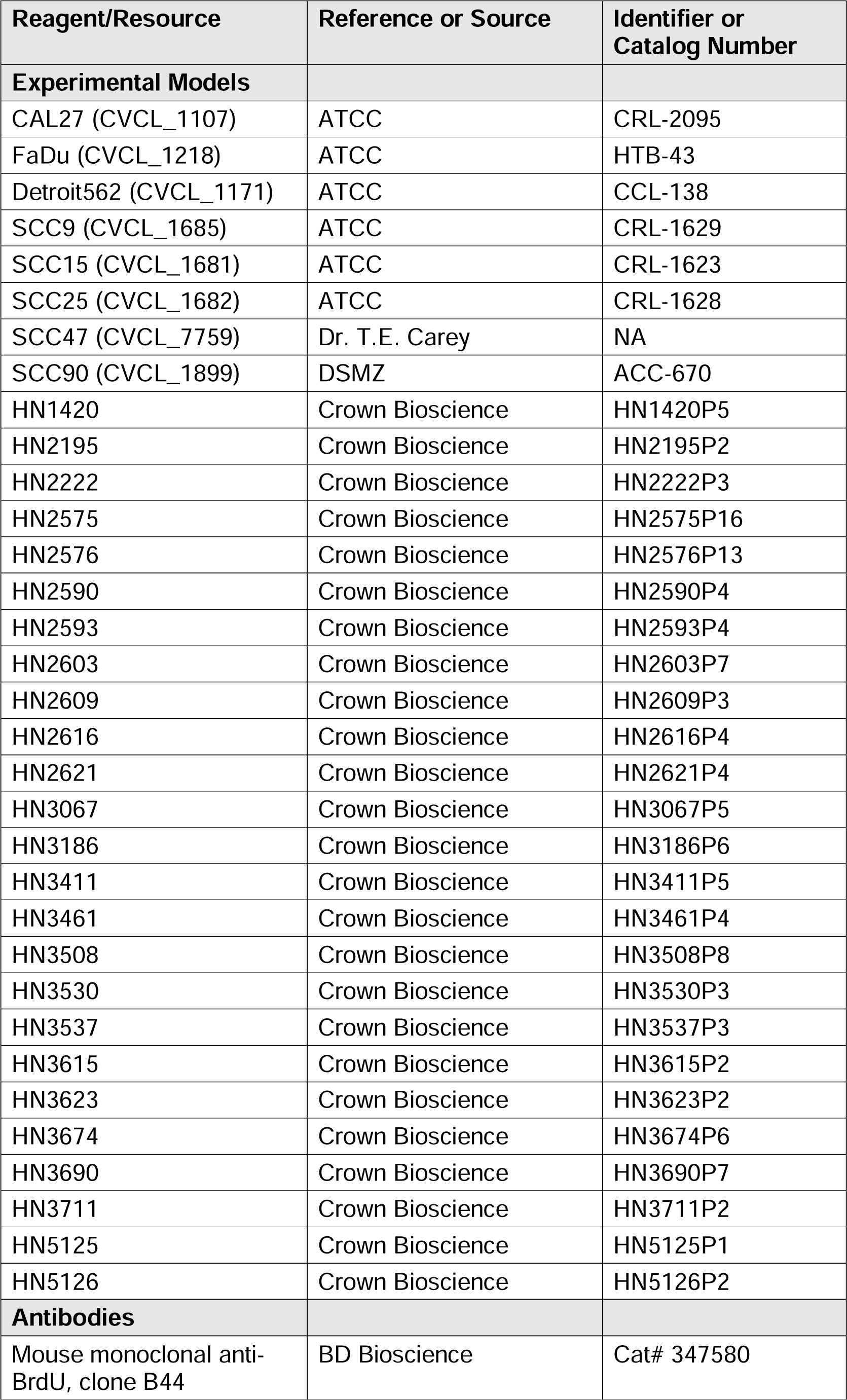
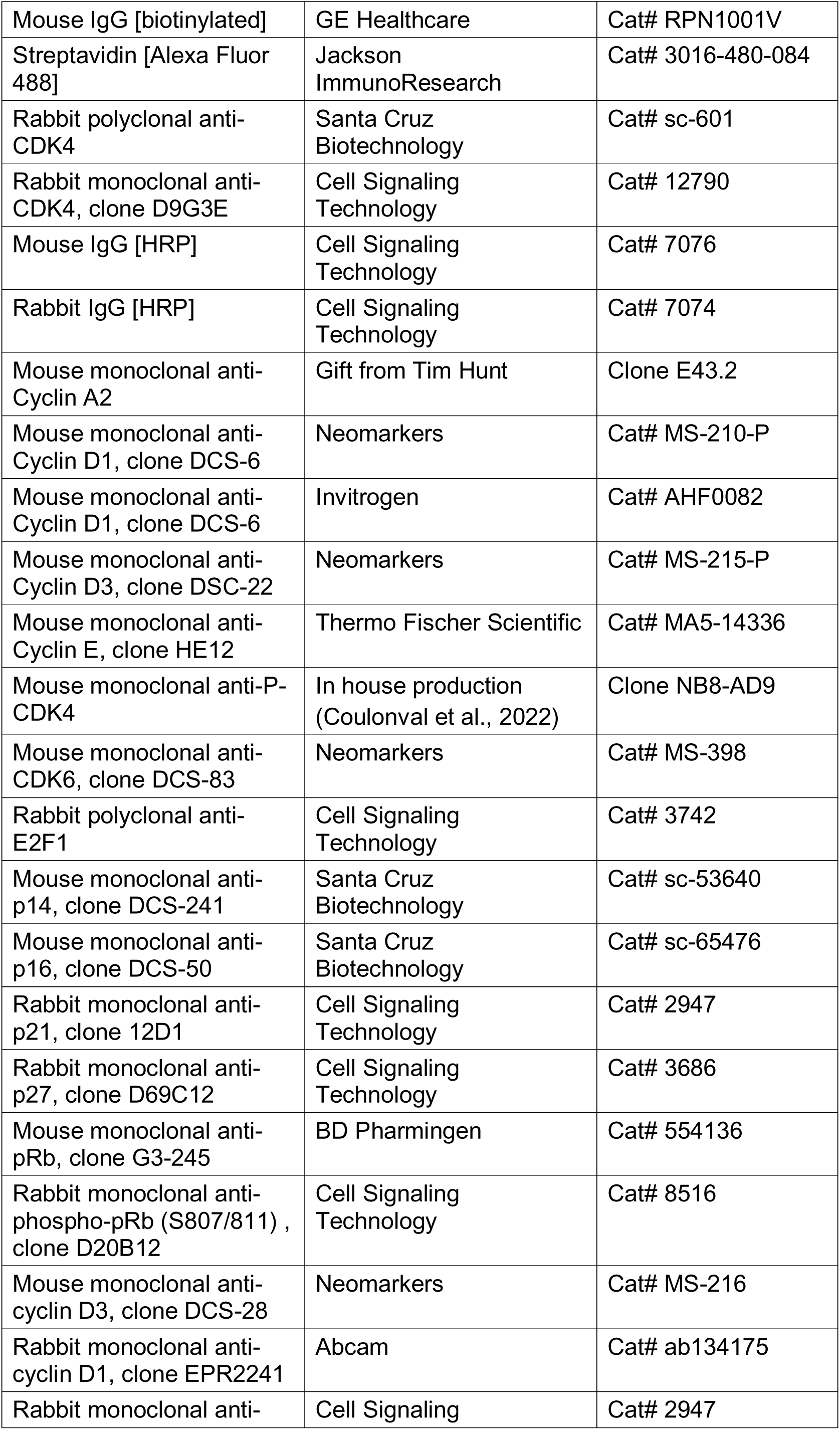
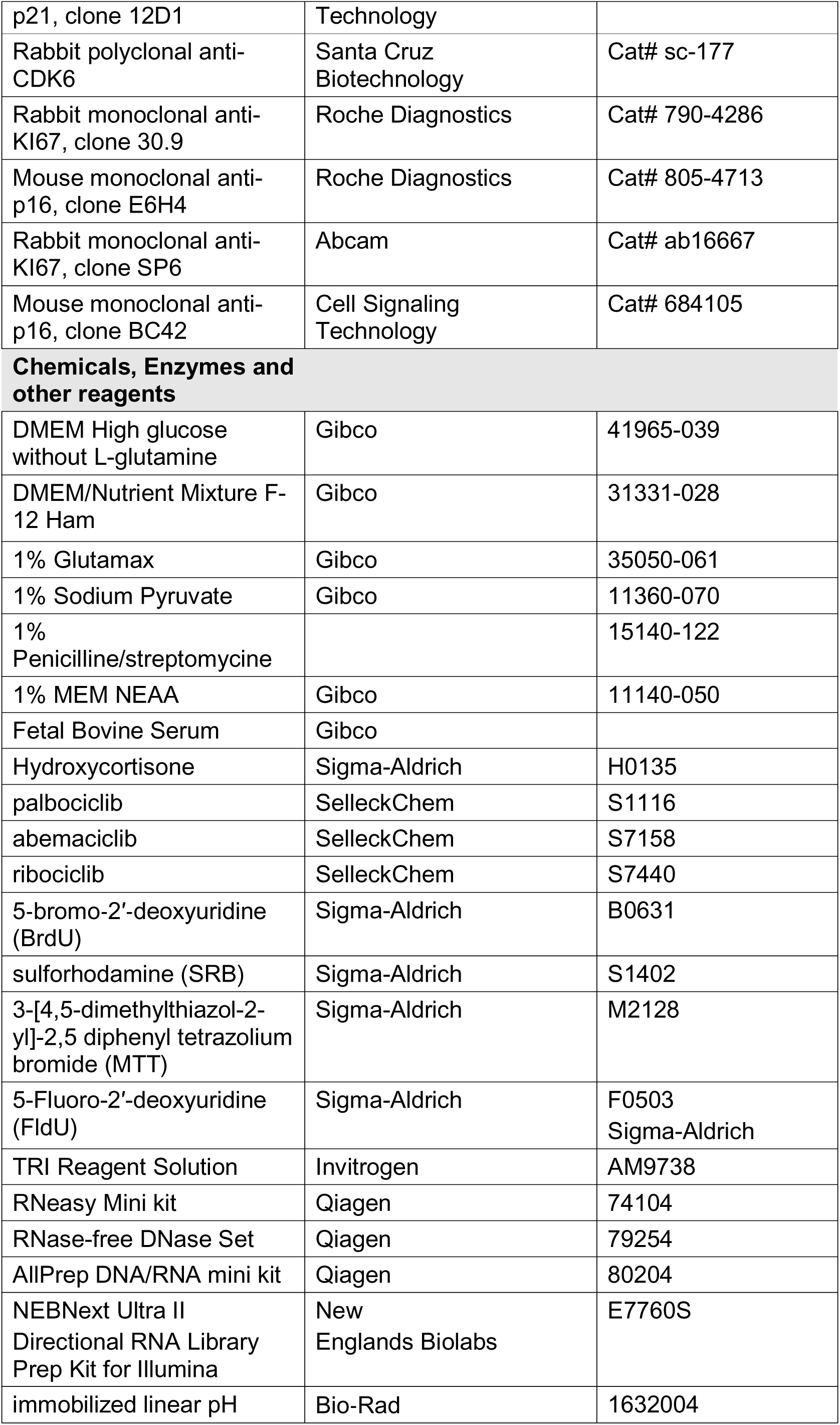
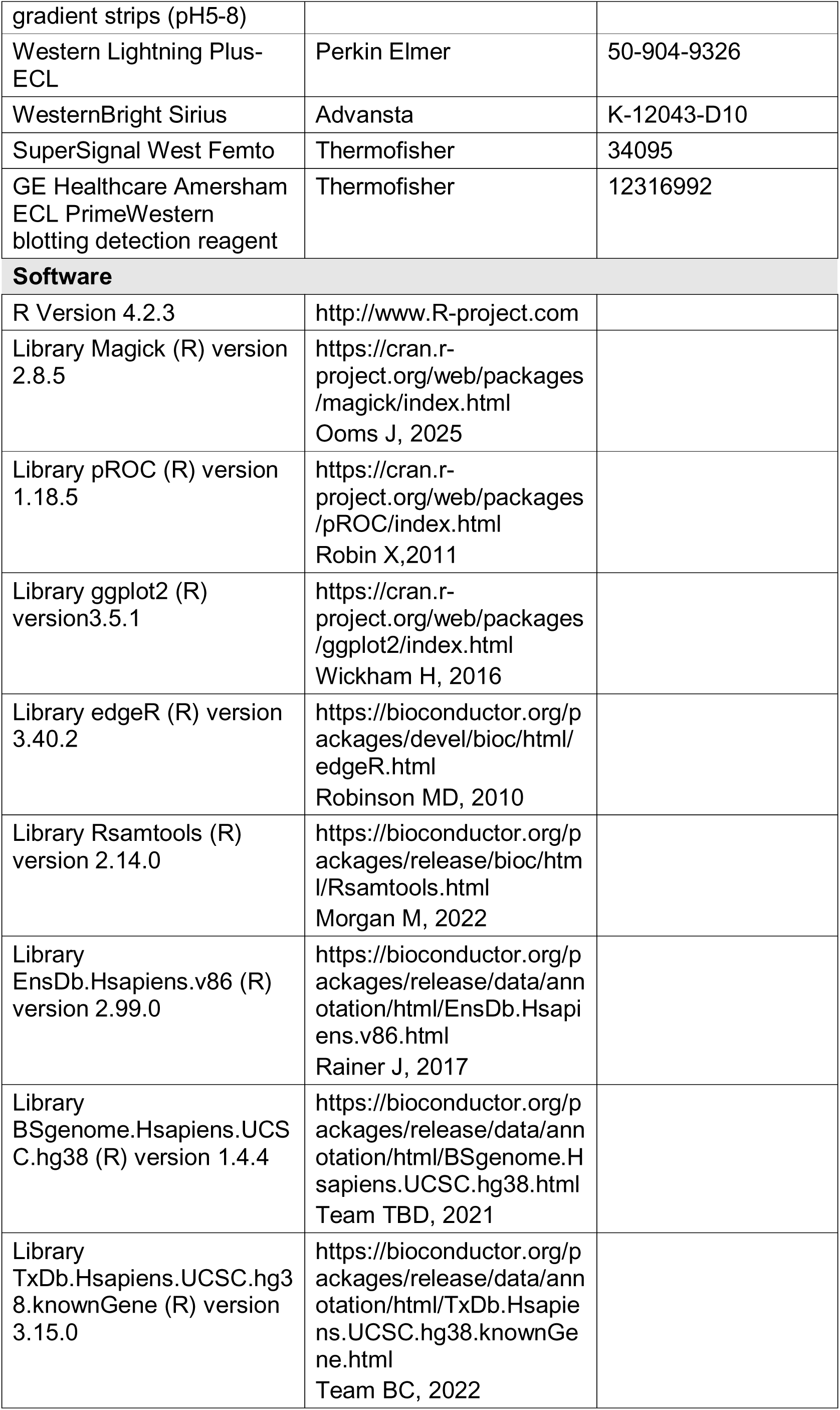
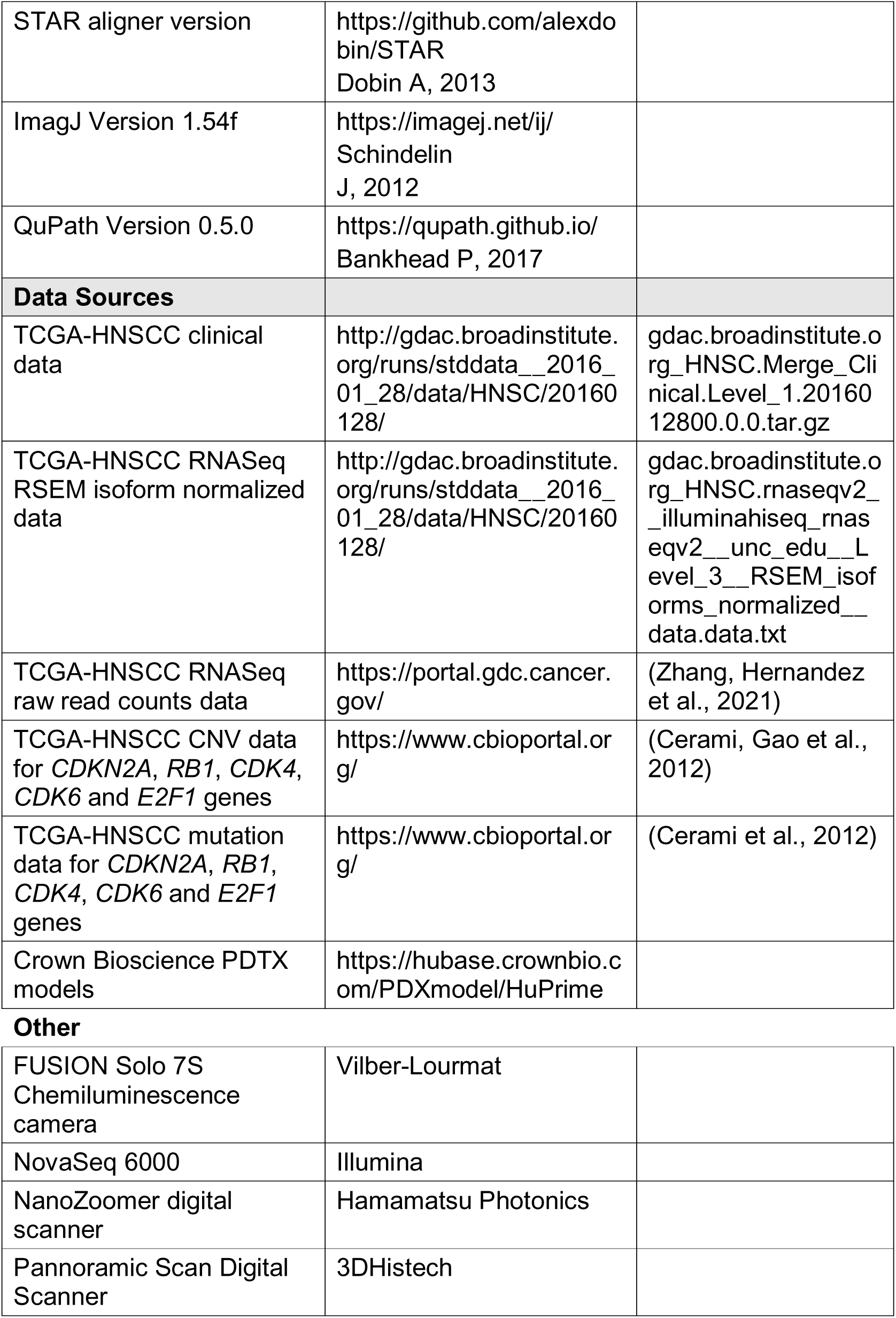

## Data Availability

Raw sequence data acquired with the RNA extracted from the tumors or PDTX of the UCLouvain and of the tumors and of the IJB were deposited at the EGA database with (submission ongoing).

## Acknowledgments

We thank Vincent Vercruysse and Antonella Mendola for technical assistance, Hajar Dahou and Bousfia Fadoua for their help in the collection of the UCLouvain tumors. We also thank Pr. A. Nichols, Y. Zimmer, R. Brakenhof and S.S. Cheong for sharing lyophilized pellets of their HNSCC cells and Pr. F Johnson for sharing HNSCC cell line molecular data. Unfortunately, the proposed dried sample preparation protocol for room temperature transport was not robust enough to avoid sample degradation and phosphorylated CDK4 signal loss precluding the use of the data in this study.

## Author’s contributions

ER, JPM and PPR successfully applied for the WalInnov grant funding the main part of this work and conceived the research. AL and FL from the ULB-BRIGHTCORE facility of the ULB analyzed the RNA extracted from all tumors and from the UCLouvain PDTX by RNASeq. WQ analyzed the RNA extracted from the Crown Bioscience PDTX by RNASeq. LZ prepared and stained the histological sections of Crown Bioscience PDTX FFPE samples and acquired the images. LZ prepared the frozen samples of Crown Bioscience PDTXs. SG supervised the histological and molecular analysis of the Crown Bioscience PDTX, validated the data and coordinated the sample and data sharing. AS and JMP prepared the frozen samples of the IJB tumors. ER and JMP extracted RNA from the IJB tumors samples. AS prepared and stained the histological sections of IJB tumor FFPE samples. ER acquired, processed, analyzed and documented the images of the IJB tumors and UCLouvain PDTX sections. PD independently validated the selection of the region of interest documenting the histology of the tumors and PDTX models, validated their characteristics and their pathological relevance. LC and FS acquired and managed the clinical data of the IJB tumors. DL supervised the histological analysis of the IJB tumors and validated the data. GvC maintained and studied the UCLouvain PDTX, collected frozen and FFPE samples thereof. KC prepared protein extracts from the IJB or the UCLouvain tumors and PDTX. KC and JMP analyzed them by 2D gel electrophoresis and Western blotting. KC extracted proteins from HNSCC cell lines and analyzed them by 1D and 2D gel electrophoresis and Western blotting. ER maintained the HNSCC cell lines in culture and studied their growth and cell cycle entry. ER analyzed the RNASeq data and performed the statistical analysis of the data. ER wrote the R and Qpath scripts used for the analysis and documentation of the data. ER coordinated the project execution. ER drew all figures of the article except Figure 3 drawn by KC and wrote the original draft of the article, which was further reviewed by JPM, XB and PPR. JPM, SG, XB and PPR supervised the study.

## The paper explained

### Problem

HNSCC is the eight most common cancer, worldwide. Patients with early disease are generally treated with single surgery or radiotherapy with a cure rate ranging between 70 to 90%. Patients with advanced disease receive multimodal treatment (surgery followed by chemo-radiation or primary chemo-radiation) with a cure rate dropping to 50%. New treatments are thus desirable. The HNSCC molecular landscape and preclinical studies suggested that the CDK4/6 inhibitory drugs used to advantageously treat advanced breast cancer may be extended to HNSCC. However, the Palatinus and NCT021034 trial results were disappointing. A better understanding of the intrinsic or acquired resistance of HNSCC to these drugs is thus required to improve their use including in combination treatments and to identify patients likely to benefit from their inclusion in their treatment or the way they should be treated.

### Results

The phosphorylation of CDK4 and CDK6 is required for the enzymes to be active. In contrast to the constitutive phosphorylation of CDK6, the CDK4 phosphorylation level is variable in cell models. CDK4 phosphorylation is absent in quiescent cells and maximal when cells are committed to execute a full cell cycle. We report here that the level of CDK4 phosphorylation is variable in HNSCC and their models and partially correlates to the IHC detection of the p16 protein together with a high proliferation index (as determined by a high proportion of Ki67- positive cells). CDK4 phosphorylation is paradoxically absent in some tumors or models with active proliferation. These models are intrinsically resistant to a treatment with CDK4/6 inhibitors, both *in vitro* and *in vivo*. Because of the requirement of CDK4 phosphorylation for the enzyme activity, this lack of CDK4 phosphorylation in proliferative tumors is the best biomarker of their intrinsic resistance to CDK4/6 inhibitors. On the other hand, some of the models in which CDK4 phosphorylation was detected and that were sensitive to CDK4/6 inhibitors *in vitro*, continued to grow *in vivo* despite treatment. CDK4 phosphorylation does thus not inform on the HNSCC capacity to acquire resistance to CDK4/6 inhibitors. Molecularly, lack of phosphorylated CDK4 was associated with defective pRb protein (due either to HPV infection, *Rb1* gene mutation or low expression) or to high expression of either *CCNE1* (leading to higher CDK2 activity and inactivation of the pRb protein) or *E2F1* genes. This lack of phosphorylated CDK4 was also associated with high expression of the p16 protein. Moreover, we provided genetic evidence that the p16 protein plays a key role in determining the CDK4 phosphorylation level. However, p16 staining alone did not faithfully identify tumors or models lacking phosphorylated CDK4 in contrast to a gene expression-based tool developed to predict the CDK4 phosphorylation status of breast tumors. Provided that the tool is adapted to HNSCC tumors by determining the *CDKN2A* gene mutation status and correcting the *CDKN2A* gene expression value by the contribution of the p16-coding mRNA, it robustly and reproducibly predicted whether or not CDK4 is phosphorylated in HNSCC.

### Impact

Since phosphorylated CDK4, the key target of CDK4/6 inhibitors is detected in the majority of HNSCC, and because their models are sensitive to these drugs at least *in vitro* when CDK4 is phosphorylated, we confirm here that their use to treat this disease is feasible and molecularly sounded. Drug resistance is a complex issue. As HNSCC and their models wherein CDK4 phosphorylation is undetectable are intrinsically resistant to the CDK4/6 inhibitors because their target is absent, our work simplifies the drug resistant issue by distinguishing intrinsically resistant tumors from the ones with acquired resistance. As the robust and efficient gene expression-based CDK4 profile prediction tool described in this work identifies the intrinsically resistant tumors and models, it will guide the appropriate way to use the CDK4/6 inhibitors to treat HNSCC. Indeed, patients with predicted phosphorylation of CDK4 are likely to benefit from a treatment with CDK4/6 inhibitor directly targeting the malignant cells provided that companion drugs are discovered and validated in order to avoid or delay drug resistance acquisition often observed following CDK4/6 inhibitor monotherapy. As MEK and CDK2 inhibitors are promising candidates as companion drug, the use of the prediction tool described in this work may increase the chance of success of trials testing the efficacy of their combination with CDK4/6 inhibitors.

On the other hand, by identifying the tumors intrinsically resistant to CDK4/6 inhibitor, the prediction tool will increase the chance of success of an alternative use of short-lived CDK4/6 inhibitors such as trilaciclib. By temporarily and reversibly blocking the cell cycle entry specifically of normal cells, this drug allows to increase the dose of chemotherapy and therefore enhance their efficacy as non-cycling cells are protected from the toxic effects of these agents.

## Legend to the EV figures

**Figure EV1.**
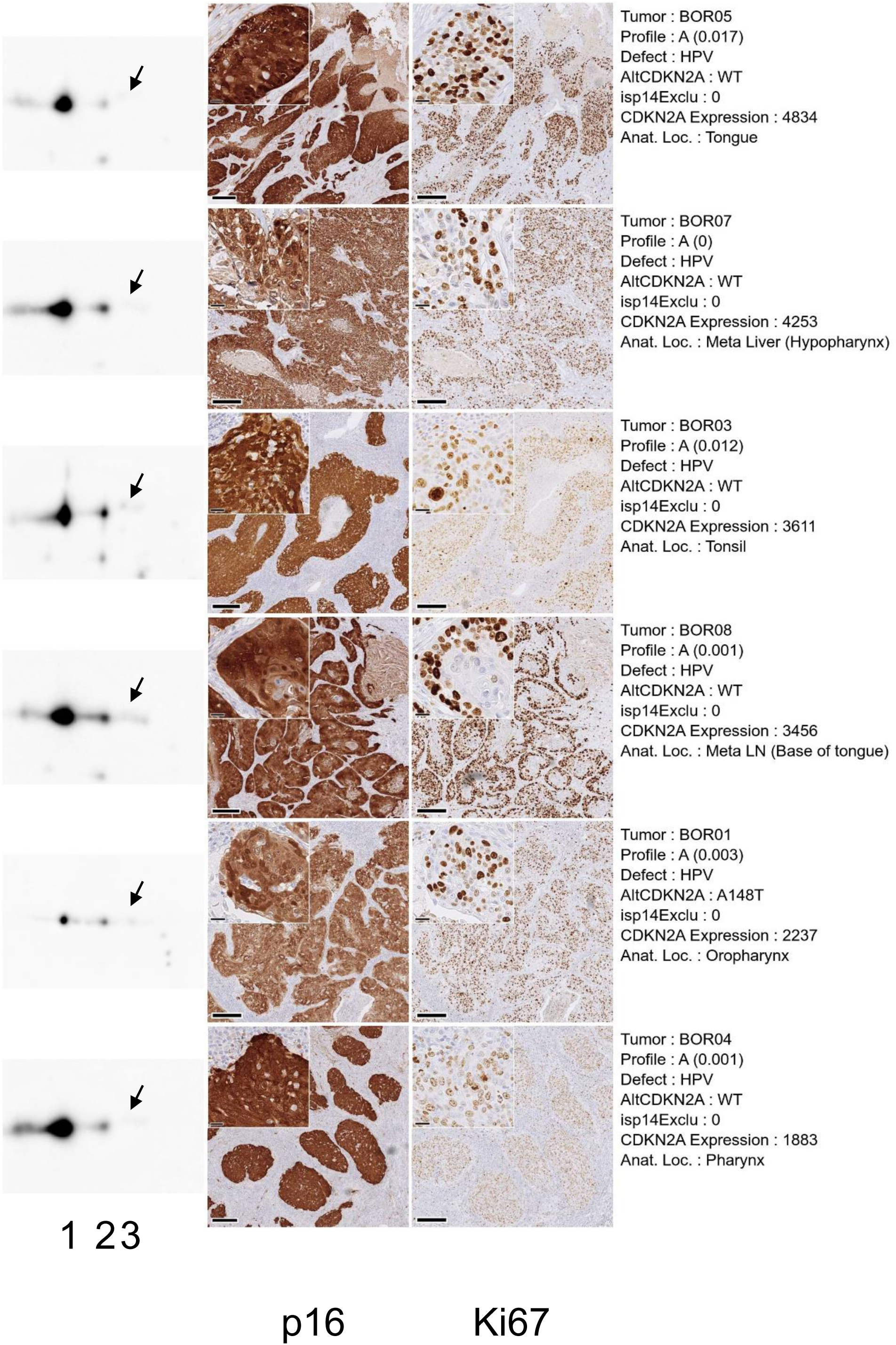
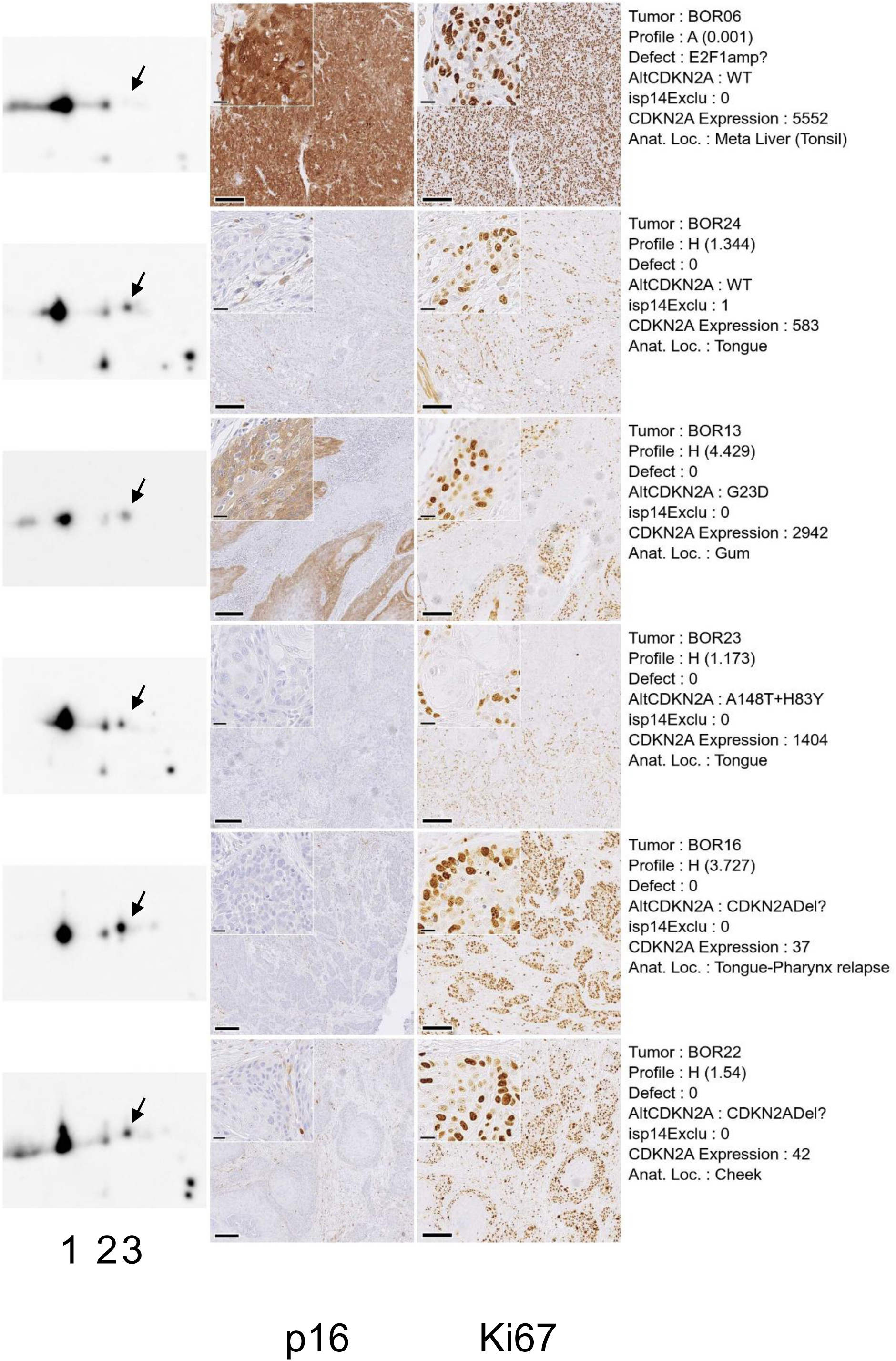
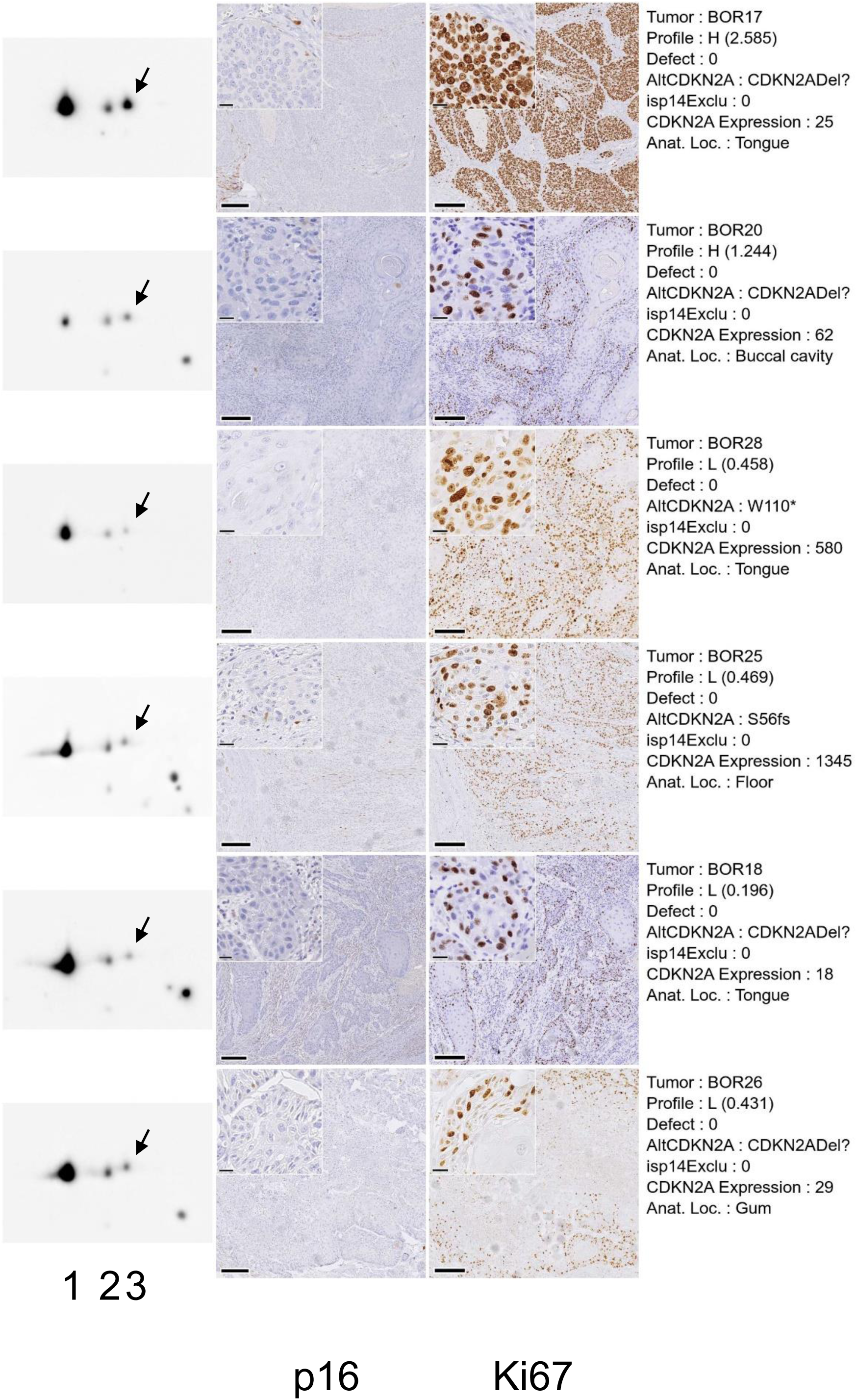
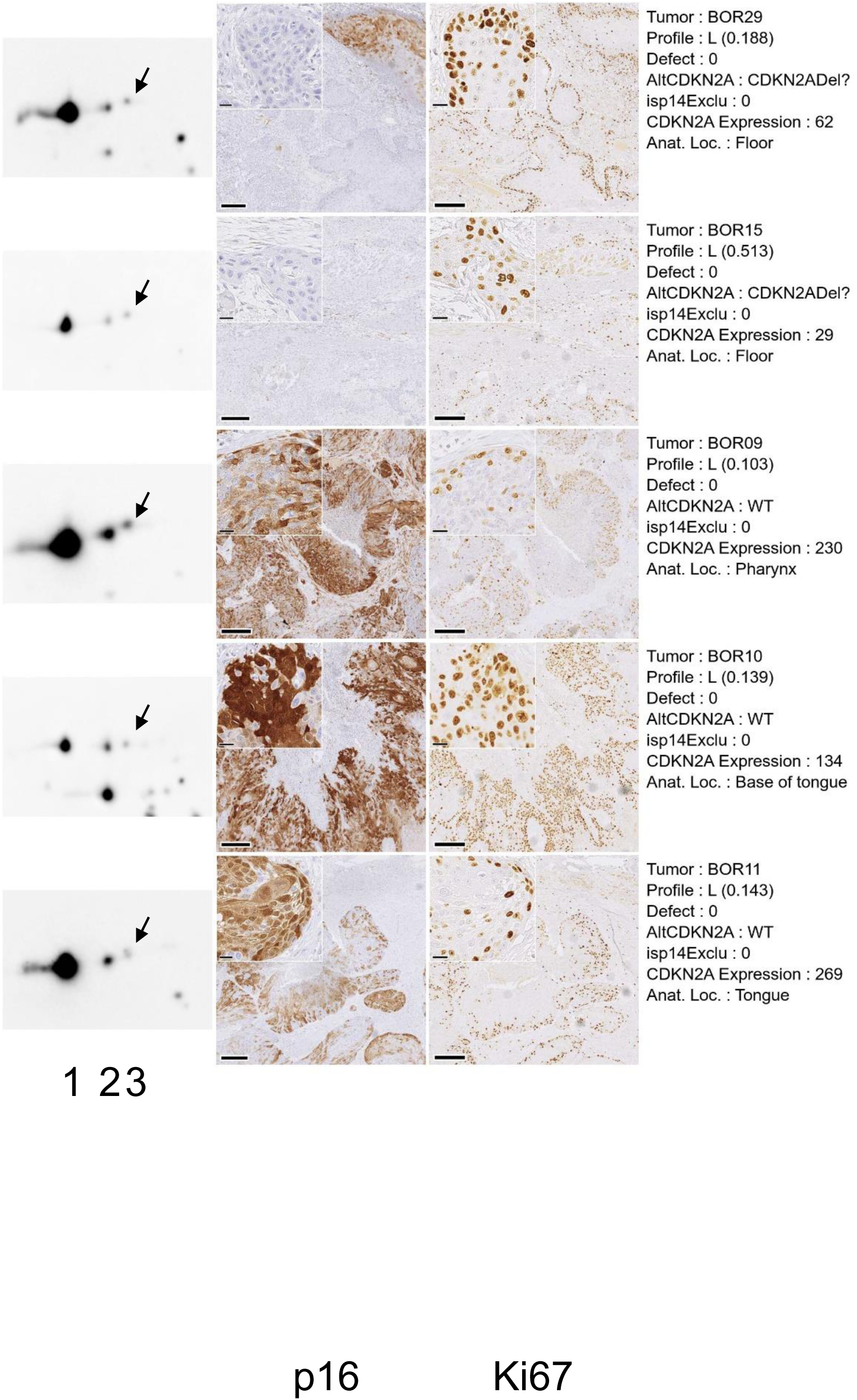
CDK4 profiles of the HNSCC tumors of the IJB cohort. Total proteins extracted from the tumors were separated by 2D electrophoresis and revealed by Western blotting with an anti-CDK4 antibody. Chemiluminescence images of the Western blots acquired with a Vilber-Lourmat Solo7S camera are shown on the left part of the figure. They were scaled to get a constant distance between spot1 (native CDK4) and the first modified form of CDK4 (spot2) measured with the Qpath software. A 300 x 200 pixels image centered on the spot2 center of mass coordinates was extracted from the whole scaled image with the magick library in R. Phosphorylated CDK4 is indicated by an arrow. A single protein extract was analyzed with a single 2D gel electrophoresis for most tumors. Due to quality issues, a second protein preparation was analyzed for the tumors BOR01, BOR03, BOR10, BOR15, BOR25, BOR18, BOR20, BOR22, BOR26 and BOR28. For all tumors except the tumors BOR03, BOR16 and BOR20 (for which only non-consecutive sections were available), 5 µm-consecutive FFPE tissue sections were stained at the pathology Department of the IJB on a fully automated BenchMark Ultra IHC system (Ventana, Roche Diagnostics, Basel, Switzerland) with the UltraView Universal DAB Detection Kit (Roche Diagnostics), using standard routine protocols. Roche monoclonal antibodies were used for primary Ki67 (clone 30.9) or p16 (clone E6H4) detection. Pictures were acquired with a NanoZoomer digital scanner (Hamamatsu Photonics, Hamamatsu, Japan) at 40× magnification. Representative regions of interest (1400 x 1400 µm images) of p16 and Ki67 IHC staining of tumors included in the IJB independent validation cohort are displayed in the middle panels as indicated in the lowest line of the figure. The scale bar on the bottom left side of the images indicates 200 µm. A 140 x 140 µm zoomed area selected in the initial region of interest is inserted on the top left part of each image. The scale bar in the bottom left of the inset indicates 20 µm. The Ki67 and p16 IHC images were aligned with the Interactive Alignment tool of the QPath software. A single Ki67 and p16 IHC was prepared for each tumor. The identities and characteristics of the tumors are indicated on the right panel. The alterations of the *CDKN2A* locus are reported in the line noted AltCDKN2A. Frame-shift mutants (indels) are noted with the amino acid affected, its position and the code fs. Mutants wherein a codon is converted to a termination codon are noted with the amino acid affected, its position and the symbol *. Samples with *CDKN2A* locus deletion or methylation are noted “CDKN2Adel?”.

**Figure EV2.**
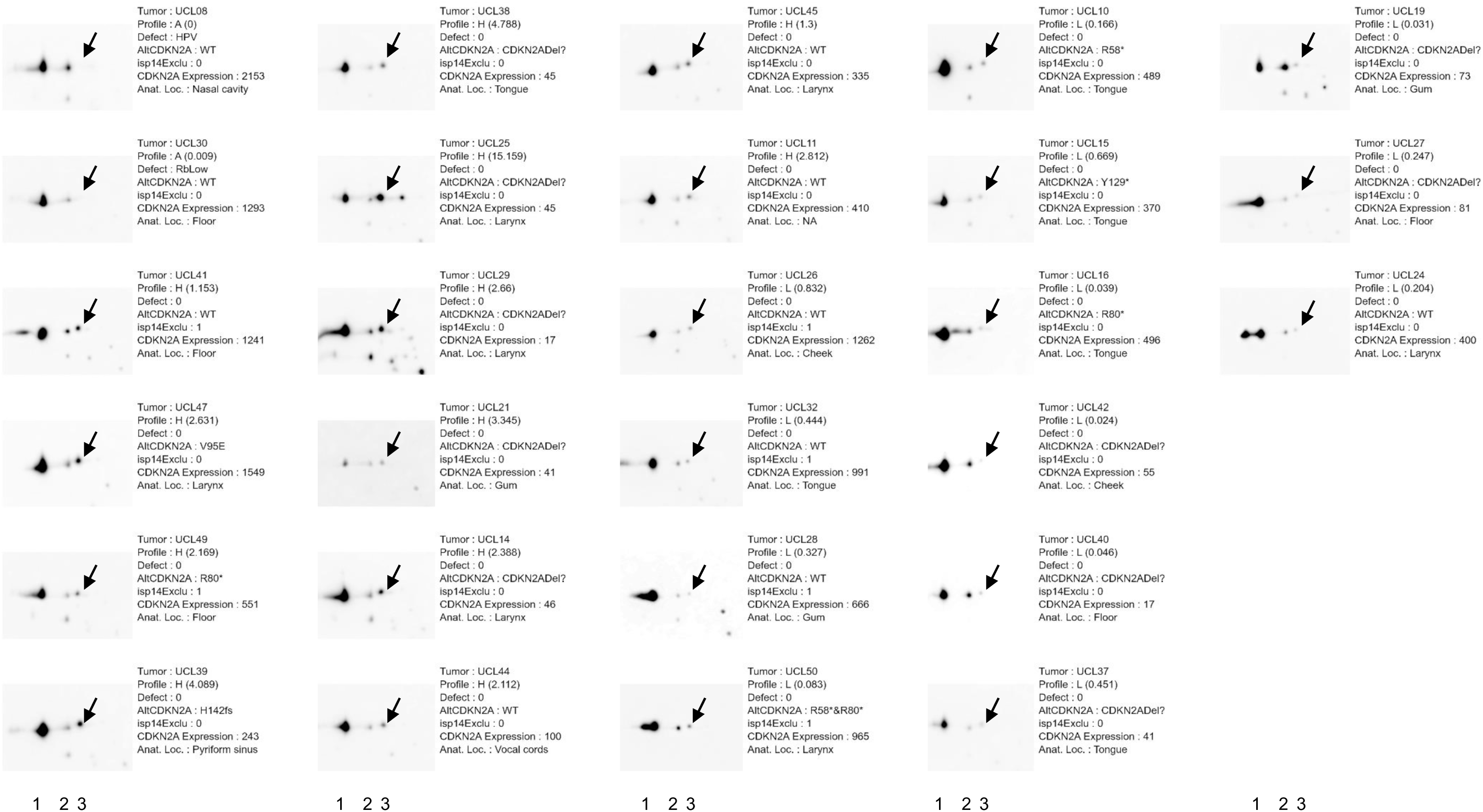
CDK4 profiles of the HNSCC tumors of the UCLouvain cohort. Total proteins extracted from the tumors were separated by 2D electrophoresis and revealed by Western blotting with an anti-CDK4 antibody. Chemiluminescence images of the Western blots acquired with a Vilber-Lourmat Solo7S camera are shown on the left of the figure. They were scaled to get a constant distance between spot1 (native CDK4) and the first modified form of CDK4 (spot2) measured with the Qpath software. A 300 x 200 pixels image centered on the spot2 center of mass coordinates was extracted from the whole scaled image with the magick library in R. Phosphorylated CDK4 is indicated by an arrow. A single protein preparation was used for tumors UCL32, UCL37, UCL38, UCL39, UCL40, UCL41, UCL42, UCL44 and UCL50. Due to quality issues, a second protein extraction was run for the other tumors. A third protein extraction was required for the tumor UCL19. The identities and characteristics of the tumors are indicated on the right of the profiles. The alterations of the *CDKN2A* locus are reported in the line noted AltCDKN2A. Frame-shift mutants (indels) are noted with the amino acid affected, its position and the code fs. Mutants wherein a codon is converted to a termination codon are noted with the amino acid affected, its position and the symbol *. Samples with *CDKN2A* locus deletion or methylation are noted “CDKN2Adel?”.

**Figure EV3.**
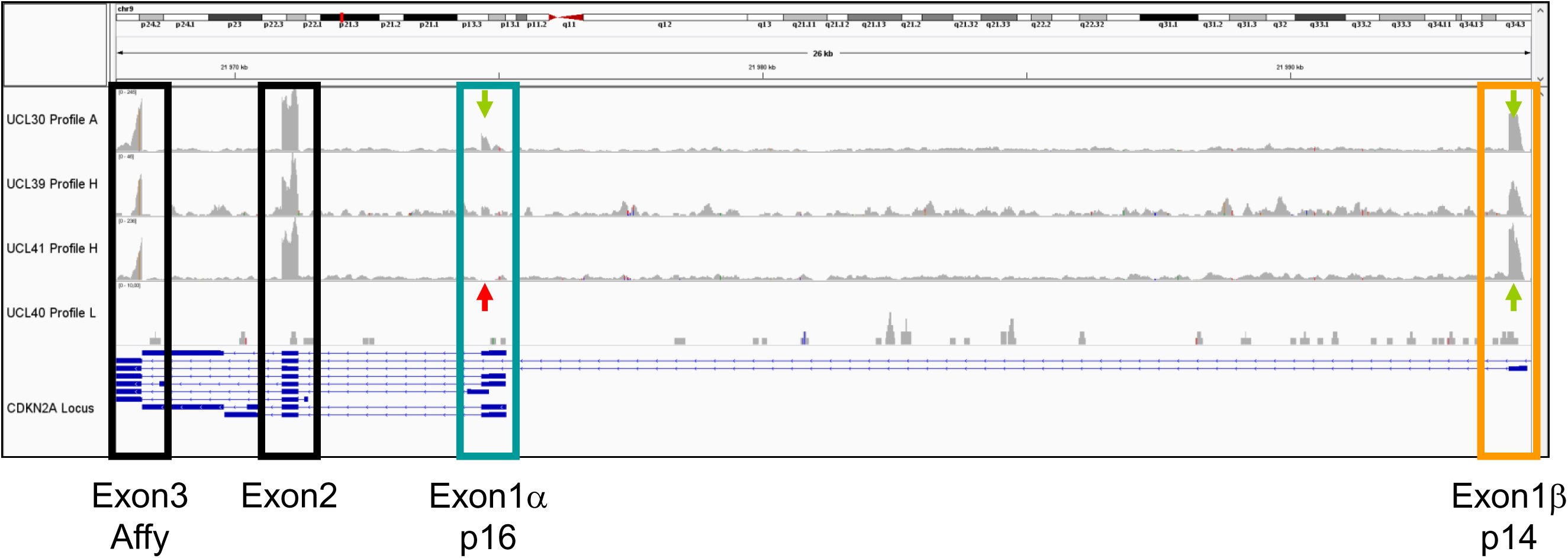
Variations of the read coverage at the *CDKN2A* locus in HNSCC tumors. RNA was extracted in parallel from the same flash frozen tumor samples as those used to extract protein and quantified by RNA sequencing. The coverage of the reads at each positions of the *CDKN2A* locus of representative tumors of the UCLouvain collection visualized with the IGV software is displayed. A single library was quantified for each tumor. The identity of the model and type of CDK4 profile are indicated on the left of the figure. The orange box identifies the exon 1β coding for the p14 protein. The green box identifies the exon 1α coding for the p16 protein. The blue box identifies the exon 3 common to the p14 and p16-coding mRNA in which the Affymetrix probes are located. Green arrows indicate strong expression at exon 1β and 1α. The red arrow at the exon 1α locus of the UCL23 sample indicates the lack of reads aligning to the exon 1α sequence.

**Figure EV4.**
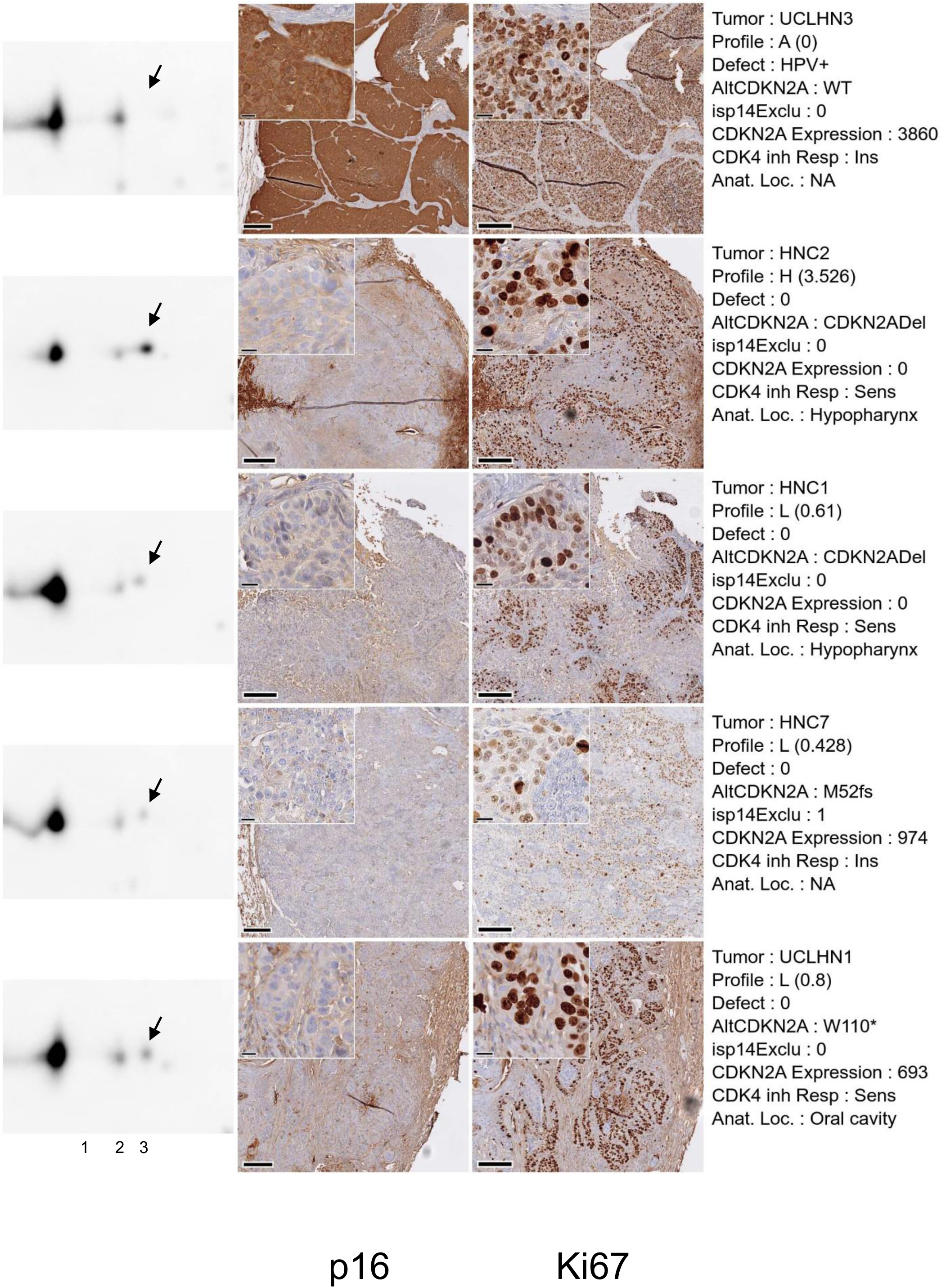
CDK4 profiles of the HNSCC PDTX models studied at the UCLouvain. Total proteins extracted from frozen samples of the selected PDTX models were separated by 2D electrophoresis and revealed by Western blotting with an anti-CDK4 antibody. Chemiluminescence images of the Western blots acquired with a Vilber-Lourmat Solo7S camera are shown on the left part of the figure. They were scaled to get a constant distance between spot1 (native CDK4) and the first modified form of CDK4 (spot2) measured with the Qpath software. A 300 x 200 pixels image centered on the spot2 center of mass coordinates was extracted from the whole scaled image with the magick library in R. Phosphorylated CDK4 is indicated by an arrow. A single protein preparation was used for each model except HNC2 due to strong contamination of the first sample with mouse tissue. A single 2D electrophoresis was run for each protein preparation. 5 µm consecutive FFPE tissue sections of the PDTX models were stained at the pathology Department of the IJB on a fully automated BenchMark Ultra IHC system (Ventana, Roche Diagnostics, Basel, Switzerland) with the UltraView Universal DAB Detection Kit (Roche Diagnostics), using standard routine protocols. Roche monoclonal antibodies were used for primary Ki67 (clone 30.9) or p16 (clone E6H4) detection. Pictures were acquired with a NanoZoomer digital scanner (Hamamatsu Photonics, Hamamatsu, Japan) at 40× magnification. Representative region of interest (1400 x 1400 µm images) of p16 and Ki67 IHC staining of the UCLouvain PDTX models are displayed in the middle panels as indicated in the lowest line of the figure. The scale bar on the bottom left side of the images indicates 200 µm. A 140 x 140 µm zoomed area selected in the initial region of interest is inserted on the top left part of each image. The scale bar in the bottom left of the inset indicates 20 µm. The Ki67 and p16 IHC images were aligned with the Interactive Alignment tool of the QPath software. A single Ki67 and p16 IHC was prepared for each model. The identities and characteristics of the models are indicated on the right panel. The alterations of the *CDKN2A* locus are reported in the line noted AltCDKN2A. Frame-shift mutants (indels) are noted with the amino acid affected, its position and the code fs. Mutants wherein a codon is converted to a termination codon are noted with the amino acid affected, its position and the symbol *. Samples with *CDKN2A* locus deletion or methylation are noted “CDKN2Adel?”.

**Figure EV5.**
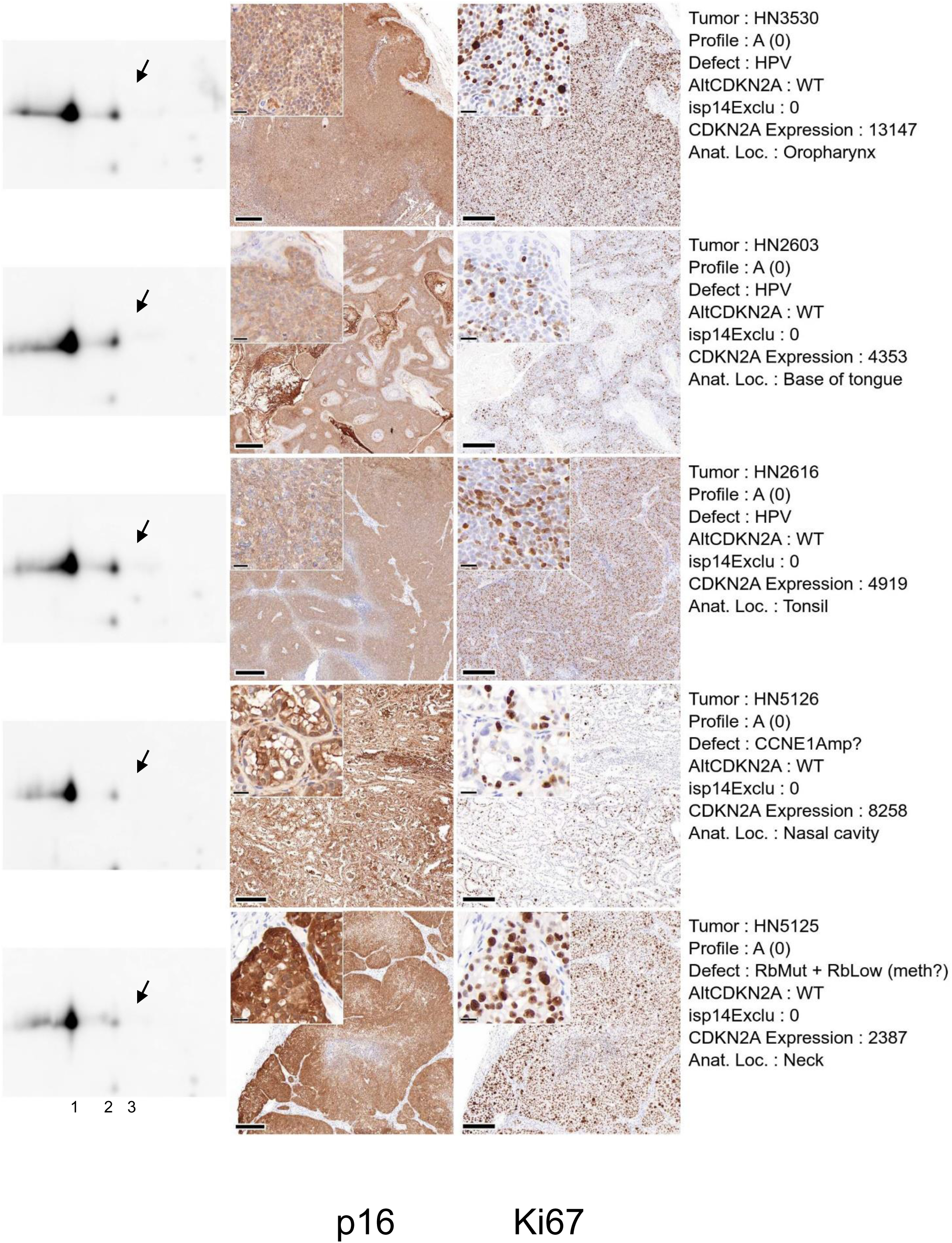
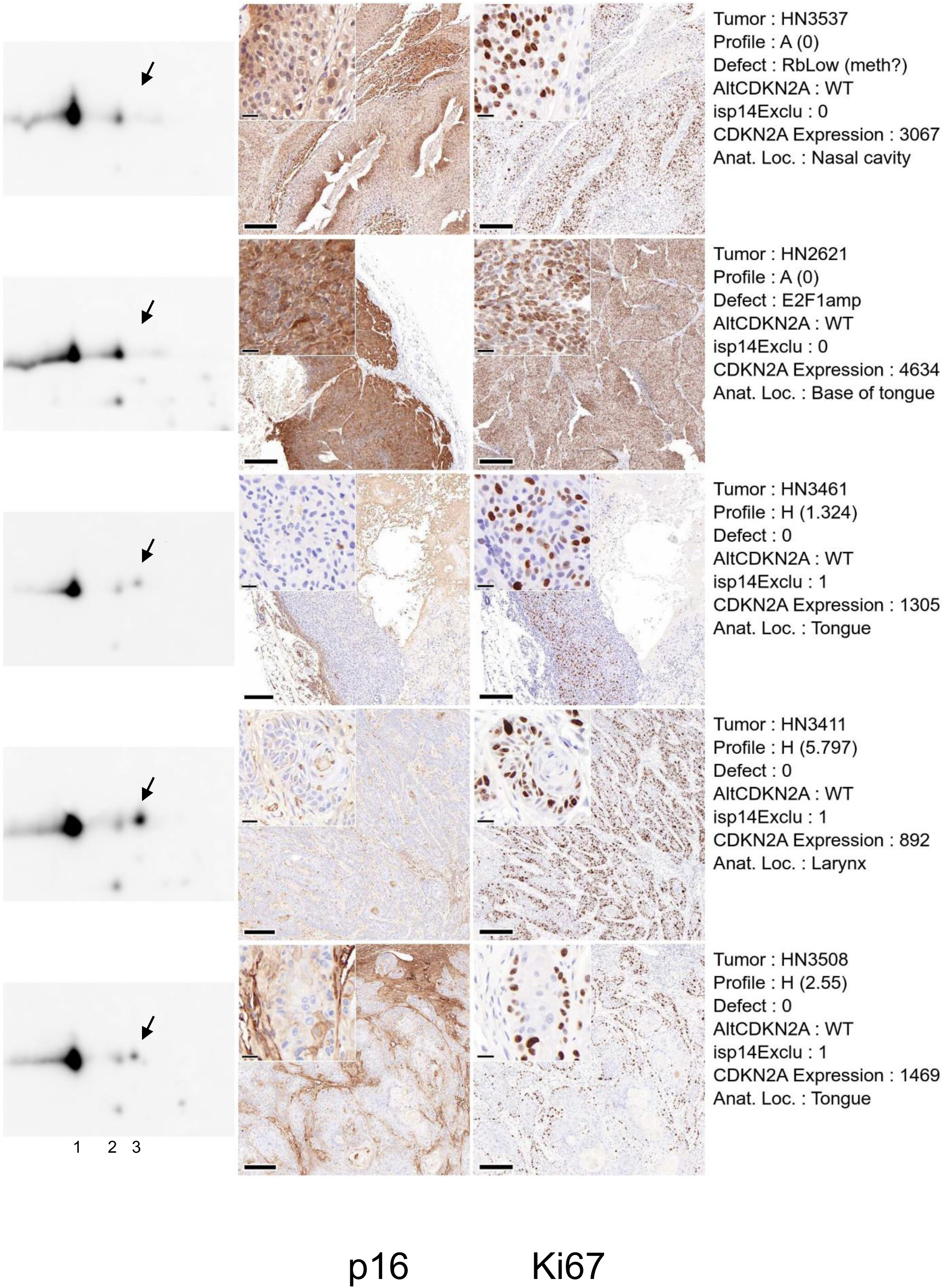
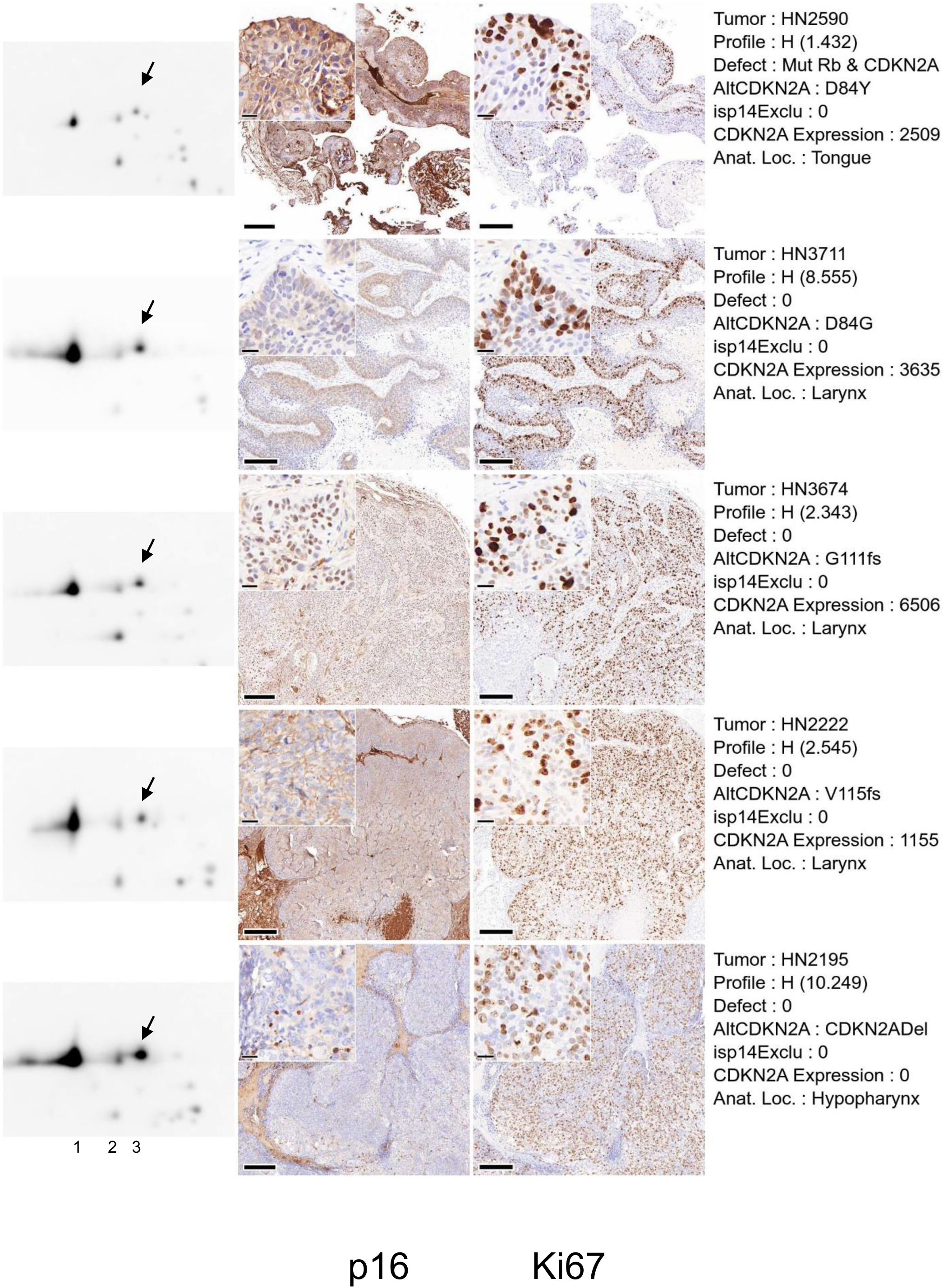
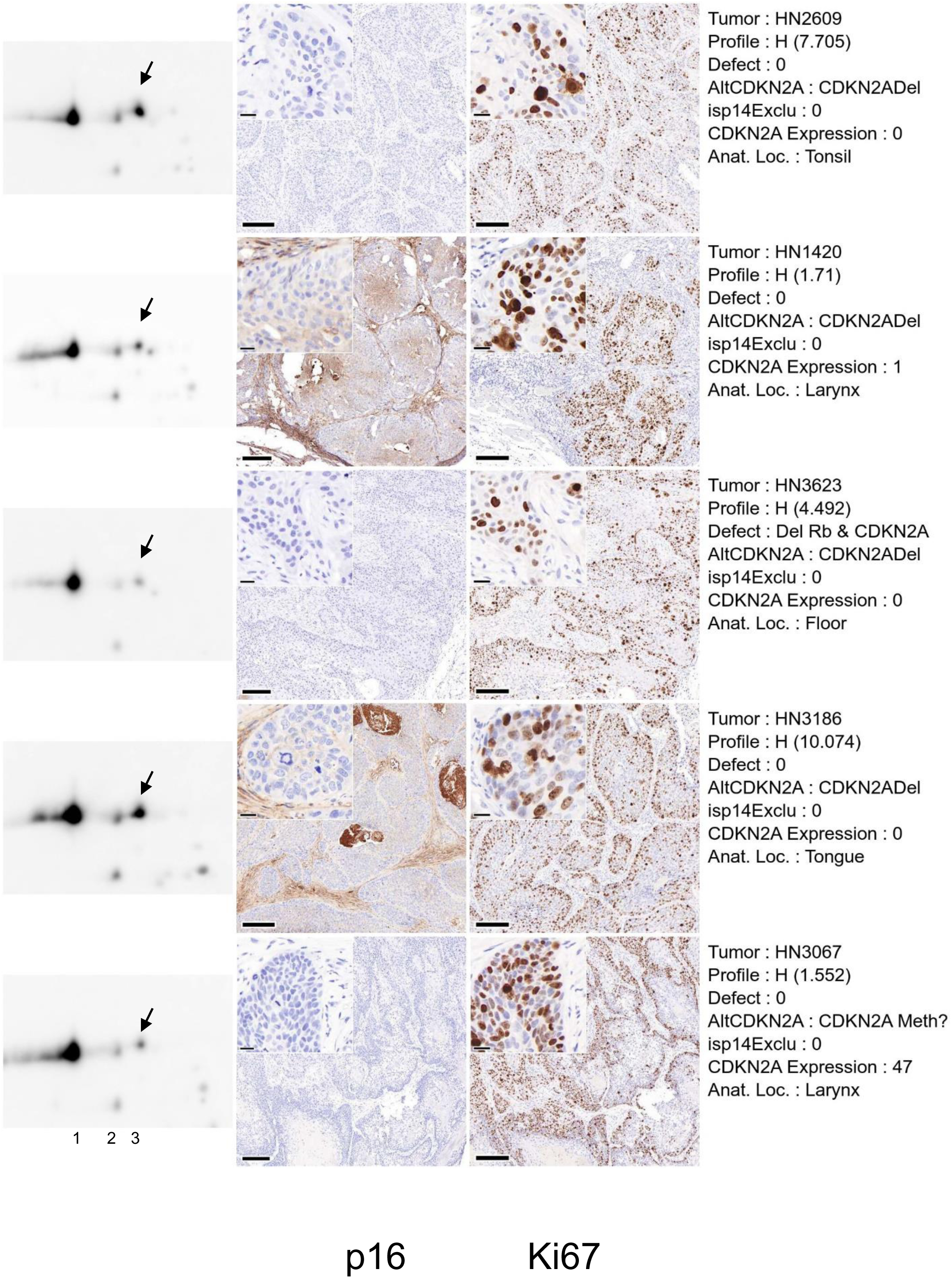
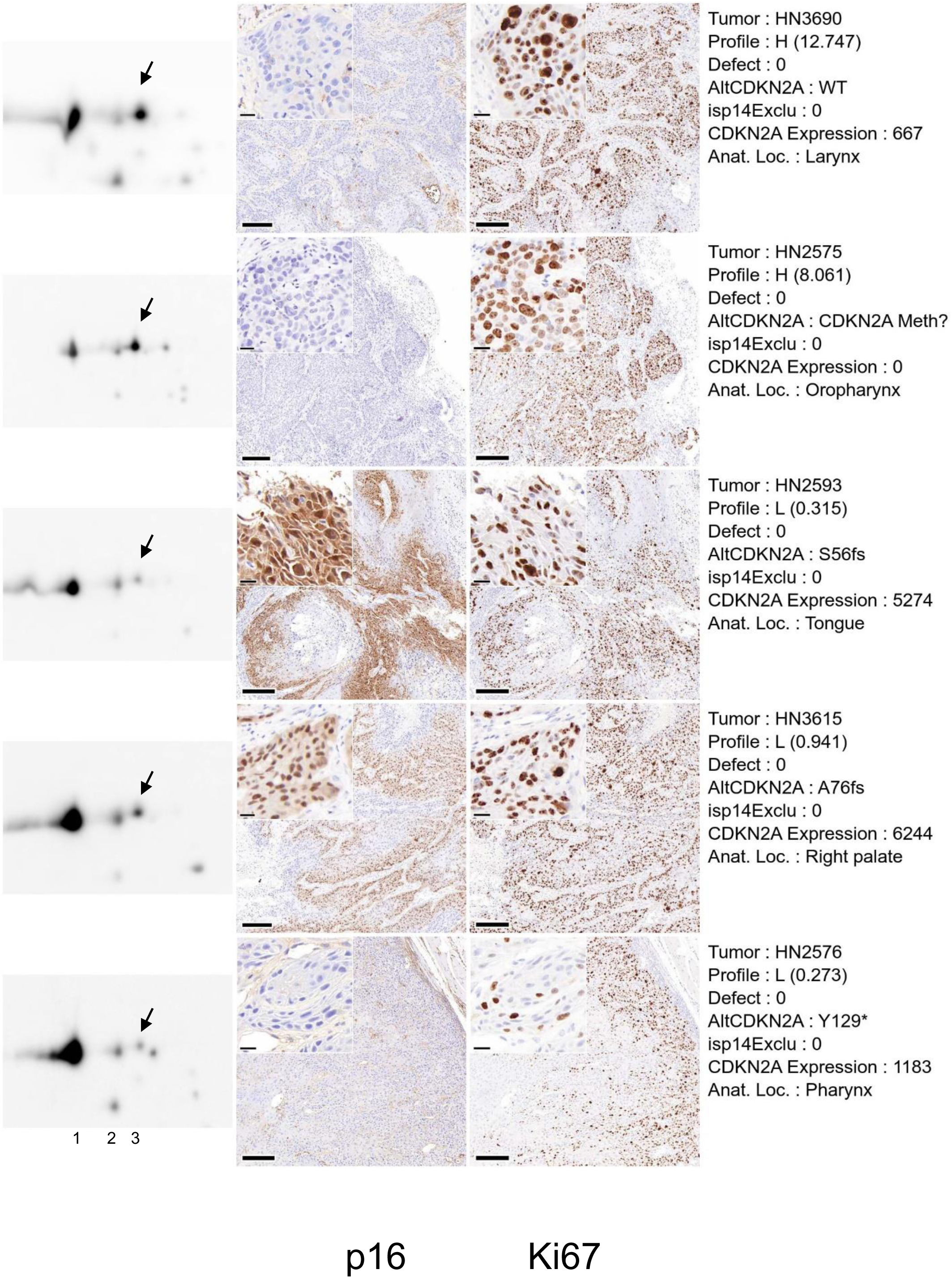
CDK4 profiles of the HNSCC tumors PDTX model selection acquired at Crown Bioscience. Total proteins extracted from frozen samples of the selected PDTX models were separated by 2D electrophoresis and revealed by Western blotting with an anti-CDK4 antibody. Chemiluminescence images of the Western blots acquired with a Vilber-Lourmat Solo7S camera are shown on the left of the figure. They were scaled to get a constant distance between spot1 (native CDK4) and the first modified form of CDK4 (spot2) measured with the Qpath software. A 300 x 200 pixels image centered on the spot2 center of mass coordinates was extracted from the whole scaled image with the magick library in R. A single protein preparation was used for each model. Due to quality issues, a second 2D gel electrophoresis was run for models HN2603, HN2621, HN2609, HN3067, HN3711, HN2576, HN2616, HN3537, HN3186. For models HN2593, HN3508, HN3530 and HN3615, three 2D gel electrophoresis were run. For all models except HN2616, HN2621 and HN1420, 5 µm consecutive sections of archival FFPE tissue of the models included in the independent validation cohort of Crown Bioscience PDTX models were available. Non-consecutive sections were used for models HN2616, HN2621 and HN1420. Sections were stained at the company using a Bond RX automatic IHC/IDH system (Leica) following their routine procedure. The Abcam anti-Ki67 (ab16667) and the Cell Signaling Technology anti-p16^INK4^ (684105) antibodies were used for primary detection revealed using the Bond Polymer Refine Detection kit (DS4800, Leica). Pictures of stained slides were acquired with a Pannoramic Scan Digital Scanner (3DHistech) at 40x magnification. Representative region of interest (1400 x 1400 µm images) of p16 and Ki67 IHC staining of tumors are displayed in the middle panels as indicated. The scale bar on the bottom left side of the images indicates 200 µm. A 140 x 140 µm zoomed area selected in the initial region of interest is inserted on the top left part of each image. The scale bar in the bottom left of the inset indicates 20 µm. The Ki67 and p16 IHC images were aligned with the Interactive Alignment tool of the QPath software. A single Ki67 and p16 IHC was prepared for each model. The identities and characteristics of the models are indicated on the right panel. The alterations of the *CDKN2A* locus are reported in the line noted AltCDKN2A. Frame-shift mutants (indels) are noted with the amino acid affected, its position and the code fs. Mutants wherein a codon is converted to a termination codon are noted with the amino acid affected, its position and the symbol *. Samples with *CDKN2A* locus deletion are noted “CDKN2Adel?”.

## Appendix

## Supplementary figures & Legends to supplementary figures

**Supplementary Figure S1.**
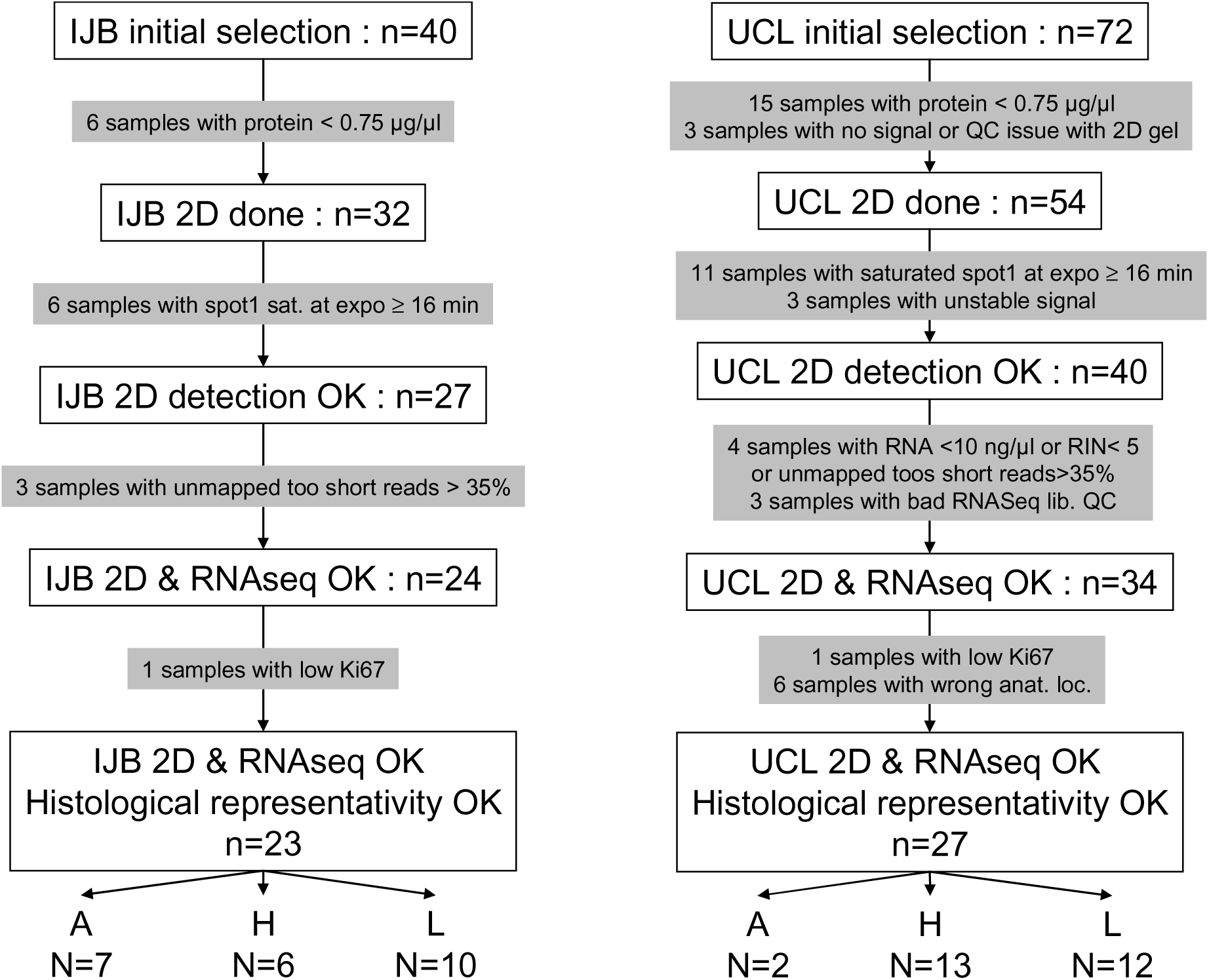
Description of the selection procedure of the HNSCC tumors of the IJB and UCLouvain cohorts. The numbers of tumors at each selection step is indicated in the open boxes. The selection criteria and numbers of excluded tumors are indicated in the grey boxes. Samples were initially selected based on the yield of proteins (more than 0.75 µg/µl) required to load the first dimension strips and reliably run a 2D gel electrophoresis. As this threshold was reduced by the later use of the WesternBright Sirius detection reagent (Advansta), the samples were selected in a second time based on the exposure times at which the native CDK4 signal is detected and saturates. Samples were excluded if no CDK4 was detected by Western blotting, or if the signal was too weak or required exposure times above 16 minutes. Several samples from the UCLouvain cohort were further excluded because the yield or quality of extracted RNA was inadequate to sequence cDNA libraries, or because RNA samples were lost or because the anatomical location of the tumor was inadequate. Finally, samples were also excluded when very few Ki67-positive cells were detected in the parallel slices stained by IHC suggesting lack of tumoral cells. IJB tumor selection chart is on the left. UCLouvain tumor selection chart is on the right.

**Supplementary Figure S2.**
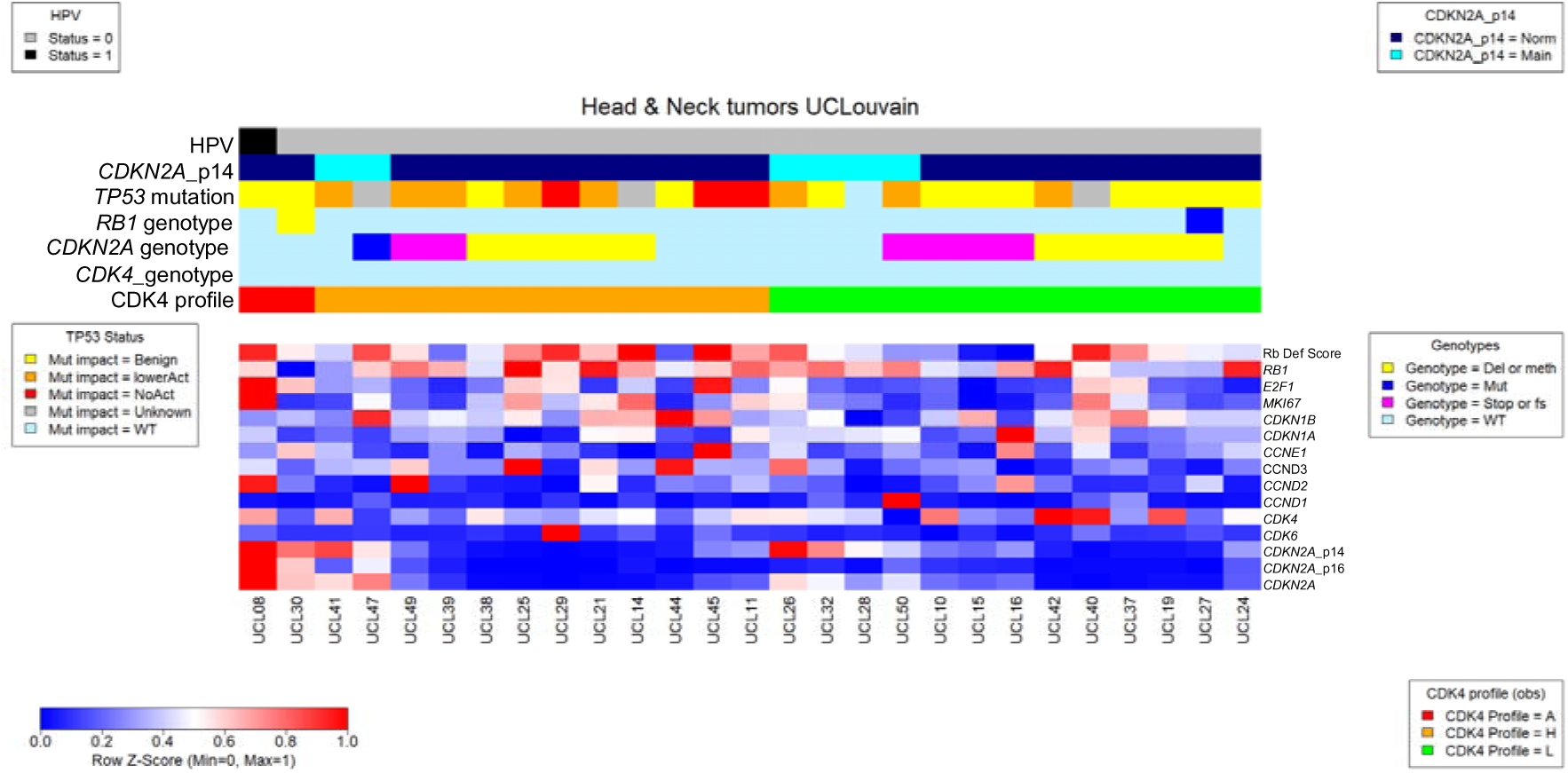
Expression of genes involved in the control or execution of the cell cycle in the human HNSCC tumors of the UCLouvain validation cohort. RNA was extracted in parallel from the same flash frozen tumor samples as those used to extract protein and quantified by RNA sequencing. Heatmaps were drawn with the heatmap.plus R package using the relative normalized expression values of the genes with the lowest value set to zero and the maximal value set to 1 as scaled in the bottom left panel of the figure. The names of the genes or measured features are displayed on the right of the heatmap. A single library was quantified for each tumor. The names of the quantified genes are indicated at the right of the heatmap. The Rb deficiency index was computed as described previously (Raspe, Coulonval et al., 2017) according to the method reported by Knudsen’s lab (Ertel, Dean et al., 2010). For the *CDKN2A* gene, the global expression of the locus (noted CDKN2A) or the expression of the locus corrected by the contributions of the mRNA coding for p16 or p14 (noted *CDKN2A*-p14 and *CDKN2A*-p16, respectively) are displayed. The HPV infection (noted HPV) status, the p14 exclusivity (noted p14), the observed *TP53*, *RB1*, *CDKN2A* and *CDK4* genotypes and the observed CDK4 profile are color coded on top of the heatmap with the features defined at the left of the color codes. The impact of the TP53 mutations is color-coded as defined in the legend on the left (NoAct = no activity due to Stop or frame shift mutations).

**Supplementary Figure S3.**
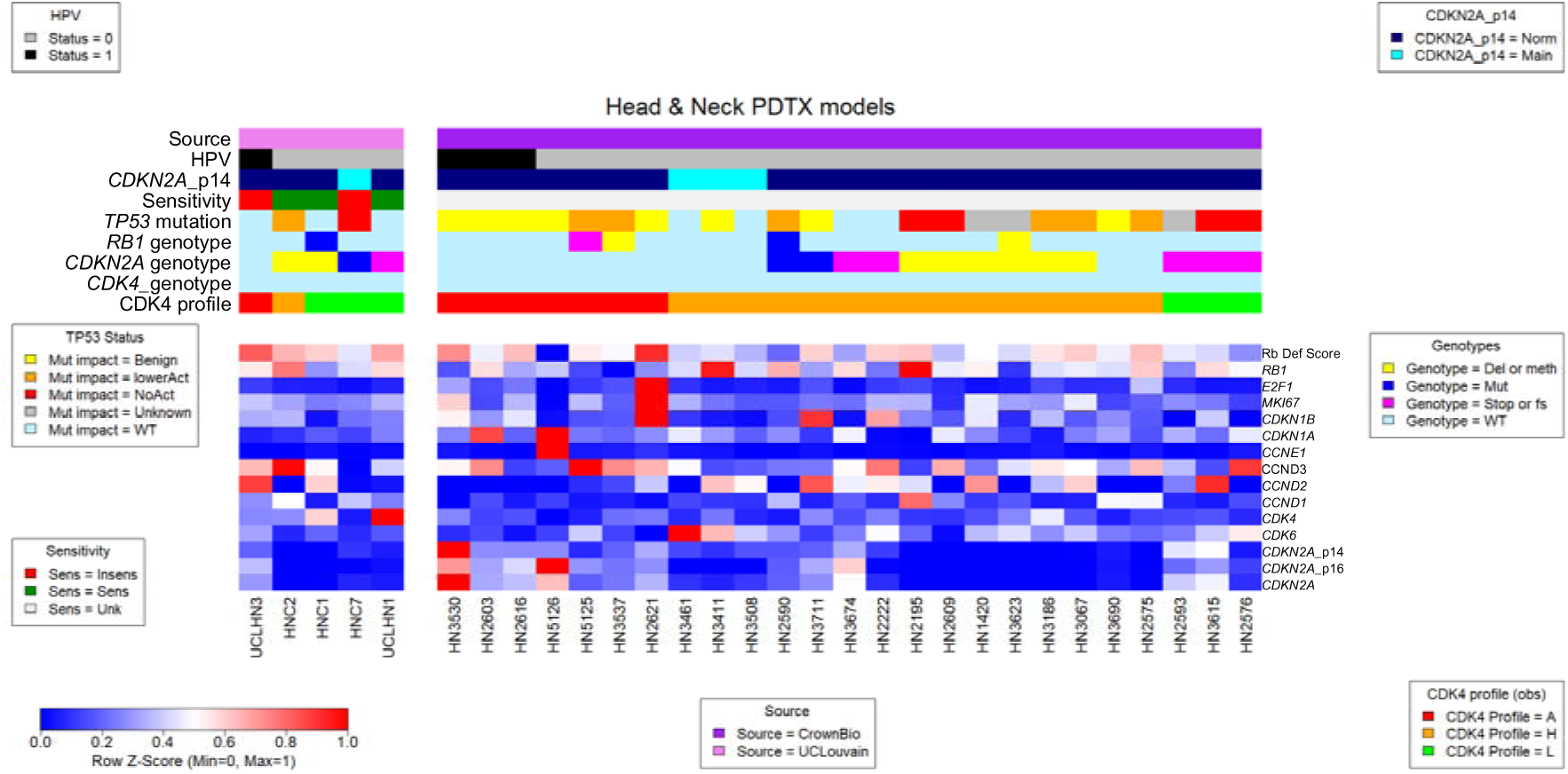
Expression of genes involved in the control or execution of the cell cycle in the human HNSCC PDTX models (UCLouvain and Crown Bioscience). RNA was extracted in parallel from the same flash frozen tumor samples as those used to extract protein and quantified by RNA sequencing. A single library was quantified for each tumor. Heatmaps were drawn with the heatmap.plus R package using the relative normalized expression values of the genes with the lowest value set to zero and the maximal value set to 1 as scaled in the bottom left panel of the figure. The names of the genes or measured features are displayed on the right of the heatmap. The Rb deficiency index was computed as described previously (Raspe et al., 2017) according to the method reported by Knudsen’s lab (Ertel et al., 2010). For the *CDKN2A* gene, the global expression of the locus (noted CDKN2A) or the expression of the locus corrected by the contributions of the mRNA coding for p16 or p14 (noted *CDKN2A*-p14 and *CDKN2A*-p16, respectively) are displayed. The HPV infection (noted HPV) status, the p14 exclusivity (noted p14), the *in vivo* CDK4/6i sensitivity status (Sensitivity), the observed *TP53*, *RB1*, *CDKN2A* and *CDK4* genotypes and the observed CDK4 profile are color-coded on top of the heatmap with the features defined at the left of the color codes. The impact of the TP53 mutations is color-coded as defined in the legend on the left (NoAct = no activity due to stop or frame shift mutations). The sensitivity codes are Insens for insensitive models, Sens for sensitive models and Unk for models not tested or for which the data are too partial to firmly establish their response to CDK4/6 inhibitors.

**Supplementary Figure S4.**
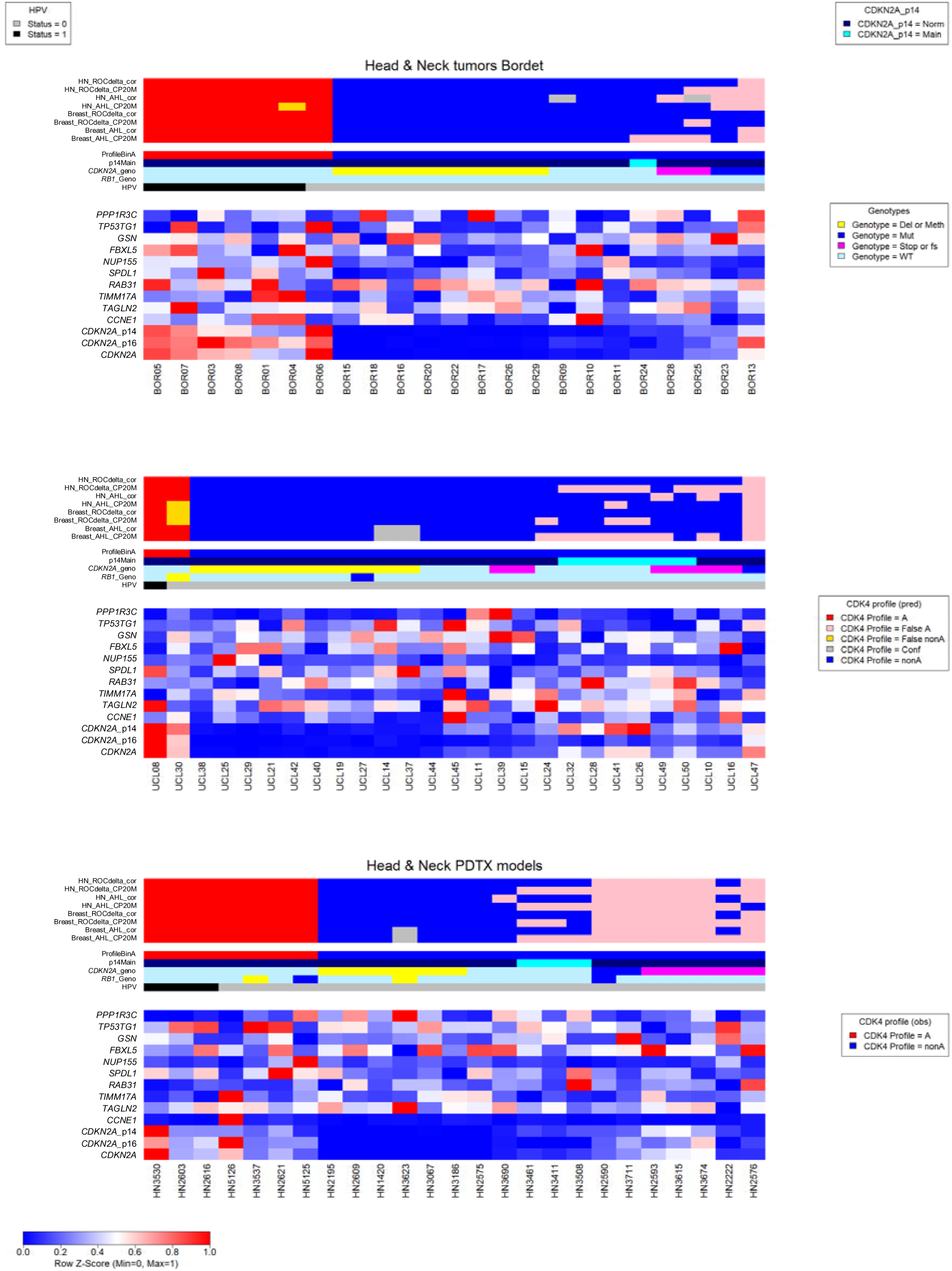
Prediction of the CDK4 profiles in HNSCC tumors and PDTX models. RNA was extracted from flash frozen tumor samples from the IJB and UCLouvain cohorts and from the Crown Bioscience HNSCC PDTX models and quantified by RNA sequencing. A single library was quantified for each tumor or model. Heatmaps were drawn with the heatmap.plus R package using the relative normalized expression values of the genes with the lowest value set to zero and the maximal value set to 1 as scaled in the bottom left panel of the figure. The names of the genes or measured features are displayed on the right of the heatmap. For the *CDKN2A* gene, the global expression of the locus (noted CDKN2A) or the expression of the locus corrected by the contributions of the mRNA coding for p16 or p14 (noted *CDKN2A*-p14 and *CDKN2A*-p16, respectively) are displayed. The HPV infection (noted HPV) status, the p14 exclusivity (noted p14Main), the observed *RB1* and *CDKN2A* genotypes and the observed CDK4 profile are color-coded on top of the heatmap with the features identified at the left of the color codes. The predictions of the CDK4 profiles with different versions of the tool are color-coded on top of these observations. From bottom to top, the tool predicts the binary (A/nonA) HNSCC CDK4 profile using breast gene expression reference if the A profile reference best correlated to the sample gene expression profile using raw expression values (tool noted Breast_AHL_CP20M) or the gene expression profile corrected for the contribution of the p16-coding mRNA to the expression of the *CDKN2A* locus (tool noted Breast_AHL_cor), the tool predicts the binary (A/nonA) HNSCC CDK4 profile using breast gene expression reference if the difference between the correlation coefficients of the comparison of the sample gene expression profile to the A and nonA references is above a threshold determined by a ROC analysis using raw expression values (tool noted Breast_ROCdelta_CP20M) or the gene expression profile corrected for the contribution of the p16-coding mRNA to the expression of the *CDKN2A* locus (tool noted Breast_ROCdelta_cor), the tool predicts the binary (A/nonA) HNSCC CDK4 profile using HNSCC gene expression if the A profile reference best correlated to the sample gene expression profile using raw expression values (tool noted HN_AHL_CP20M) or the gene expression profile corrected for the contribution of the p16-coding mRNA to the expression of the *CDKN2A* locus (tool noted HN_AHL_cor), the tool predicts the binary (A/nonA) HNSCC CDK4 profile using HNSCC gene expression reference if the difference between the correlation coefficients of the comparison of the sample gene expression profile to the A and nonA references is above a threshold determined by a ROC analysis using raw expression values (tool noted HN_ROCdelta_CP20M) or the gene expression profile corrected for the contribution of the p16-coding mRNA to the expression of the *CDKN2A* locus (tool noted HN_ROCdelta_cor). If the two highest correlation coefficients of the comparison of the sample gene expression profile are equal and include the one of the A profile reference, the profile cannot be predicted and is colored in grey (Conf for conflicting). False prediction of A profile is colored in pink while false prediction of non A profile is colored in gold. Samples were reordered according to the prediction accuracy, the *CDK2A* mutation status and the exclusivity of the p14-coding mRNA expression.

## Description of the EV datasets

### Dataset EV1. Molecular and clinical features of the final selection of HNSCC tumors from the IJB and UCLouvain cohorts

The molecular characteristics and clinical features of the tumors from the IJB and UCLouvain cohorts included in the analysis are reported respectively in the worksheet “IJB cohort” and “UCLouvain cohort”. Description of the column content is found below each table. Frame-shift mutants (indels) are noted with the amino acid affected, its position and the code fs. Mutants wherein a codon is converted to a termination codon are noted with the amino acid affected, its position and the symbol *. Samples with *CDKN2A* locus deletion are noted CDKN2Adel.

### Dataset EV2. PDTX models characteristics

The names, references, sources, clinical characteristics and molecular features of the Crown Bioscience models used in this study are reported respectively in the worksheets names “Crown Bioscience” and “UCLouvain”. Description of the column content is found below the table. The mutations described in the “Crown Bioscience” worksheet were collected on the Crown Bioscience website (https://hubase.crownbio.com/PDXmodel/HuPrime) and checked by visualization with the IGV software (https://igv.org/).

The “Selection_QC_PDTX_Crownbio” and “Selection QC_PDTX_UCLouvain” worksheets report the statistics acquired by the analysis of the samples in this work and their use to select the samples. Description of the column content is found below the table. Frame-shift mutants (indels) are noted with the amino acid affected, its position and the code fs. Mutants wherein a codon is converted to a termination codon are noted with the amino acid affected, its position and the symbol *. Samples with *CDKN2A* locus deletion are noted CDKN2Adel.

### Dataset EV3. Cell lines characteristics

The names, references, sources, characteristics and molecular features of the cell lines used in this study are reported. The composition of the culture medium used and description of the column content are indicated below the table. Frame-shift mutants (indels) are noted with the amino acid affected, its position and the code fs. Mutants wherein a codon is converted to a termination codon are noted with the amino acid affected, its position and the symbol *. Samples with *CDKN2A* locus deletion are noted CDKN2Adel.

### Dataset EV4 Selection of the HNSCC tumors from the IJB and UCLouvain collection

The “Selection IJB” and “Selection_UCLouvain” worksheets report the analyte statistics acquired during the analysis of the tumor samples in this work. The selected samples used for the final CDK4 profile analysis and prediction are identified. The reason for discarding samples are indicated. Description of the column content is found below the table.

## Supplementary tables

**Supplementary Table S1.** Selected gene expression values in HNSCC tumors models with H or L CDK4 profiles. The averages and standard deviation of the expression values of the indicated gene expression levels or of the Rb deficiency index in tumors with H (n=40) or L (n=34) are reported. A non-parametric Wilcoxon test was run in R to test the null hypothesis of equal means as the variances were not always the same and as the null hypothesis of normal distributions of the values in the H and L subsets can often be rejected. The Wilcoxon and Shapiro test p values and the variances of the values in the profile H and L subsets are reported.

|  | AVG<br>total H | SD<br>total H | AVG<br>total L | SD<br>total L | Wilcoxon test<br>pval | Shapiro<br>test<br>pval_H | Shapiro<br>test<br>pval_L | Var<br>H | Var<br>L |
| --- | --- | --- | --- | --- | --- | --- | --- | --- | --- |
| Rb_def_score | -0.1886 | 0.1494 | -0.2947 | 0.233 | 1.58E-01 | **6.20E-01** | **1.01E-01** | 0.022331 | 0.054298 |
| p16 | 106.7 | 245.2 | 34.6 | 46.1 | 9.26E-01 | 4.23E-07 | 1.93E-04 | 60108.43 | 2125.29 |
| p14 | 404.8 | 560.9 | 356.5 | 401.9 | 7.64E-01 | 1.50E-04 | 7.28E-04 | 314650.5 | 161496.5 |
| RB1 | 1776.7 | 483.9 | 1907 | 664.8 | 5.65E-01 | **1.49E-01** | 1.04E-02 | 234181.5 | 441902.1 |
| E2F1 | 288.9 | 149 | 259 | 115.6 | 4.88E-01 | 1.68E-02 | **1.09E-01** | 22210.94 | 13374.9 |
| MKI67 | 3823.2 | 1355 | 3668.8 | 1620.8 | 6.10E-01 | **6.13E-01** | **1.16E-01** | 1836094 | 2627118 |
| CDKN1B | 1306.5 | 432.6 | 973.2 | 259.2 | **3.20E-02** | **2.14E-01** | **1.35E-01** | 187157.2 | 67183.2 |
| CDKN1A | 3649 | 1662 | 4651.9 | 2081.5 | 8.92E-02 | **3.45E-01** | **1.65E-01** | 2762198 | 4332545 |
| CCNE1 | 121.9 | 69.6 | 149.5 | 76.2 | 8.92E-02 | 4.87E-04 | 1.04E-02 | 4841.497 | 5810.26 |
| CCND3 | 1013.7 | 381.5 | 852.6 | 490.4 | 5.63E-02 | **1.46E-01** | 2.41E-03 | 145575.1 | 240478.5 |
| CCND2 | 4835.1 | 4460.3 | 3365.2 | 3356.7 | 2.84E-01 | 1.79E-03 | 4.74E-05 | 19894431 | 11267633 |
| CCND1 | 5420.8 | 3374.7 | 11091.3 | 16081 | 5.48E-01 | 9.57E-03 | 1.12E-06 | 11388448 | 2.59E+08 |
| CDK4 | 1666.7 | 1155.7 | 1335.6 | 447.9 | 4.48E-01 | 4.63E-06 | **5.83E-01** | 1335661 | 200593.2 |
| CDK6 | 3473.7 | 1195.9 | 4226.4 | 2171.3 | 3.74E-01 | **4.85E-01** | **1.72E-01** | 1430281 | 4714702 |
| CDKN2A | 511.5 | 767.5 | 391 | 418.6 | 8.65E-01 | 4.30E-05 | 1.67E-03 | 589120.2 | 175216.1 |
| TAGLN2 | 7105.2 | 1751.8 | 7951.4 | 1663.3 | 1.58E-01 | **5.47E-01** | **9.98E-01** | 3068817 | 2766611 |
| TIMM17A | 628.9 | 167.1 | 625.6 | 176.2 | 8.04E-01 | **5.40E-02** | **7.02E-02** | 27927.99 | 31047.87 |
| RAB31 | 2771.1 | 1336.2 | 3581 | 1373.8 | 1.05E-01 | **2.22E-01** | **9.18E-01** | 1785338 | 1887356 |
| SPDL1 | 333.3 | 125.1 | 312.8 | 127 | 4.72E-01 | **6.94E-01** | 3.00E-03 | 15660.54 | 16123.23 |
| NUP155 | 1799.7 | 1072.4 | 1728.3 | 661.3 | 4.80E-01 | 1.17E-04 | 6.30E-04 | 1149950 | 437319.2 |
| FBXL5 | 1414.6 | 302.3 | 1479.1 | 402.9 | 6.38E-01 | **1.33E-01** | 2.92E-03 | 91403.15 | 162315.8 |
| GSN | 7976.3 | 2658.5 | 6666.4 | 2209.4 | 1.66E-01 | **2.44E-01** | **2.95E-01** | 7067765 | 4881618 |
| TP53TG1 | 117.7 | 64.4 | 98.7 | 46.1 | 4.18E-01 | **6.61E-02** | **1.61E-01** | 4143.094 | 2127.084 |
| PPP1R3C | 416.4 | 730.8 | 255.8 | 244.3 | 7.14E-01 | 5.96E-07 | 1.23E-03 | 534141.3 | 59671.68 |

**Supplementary Table S2.**
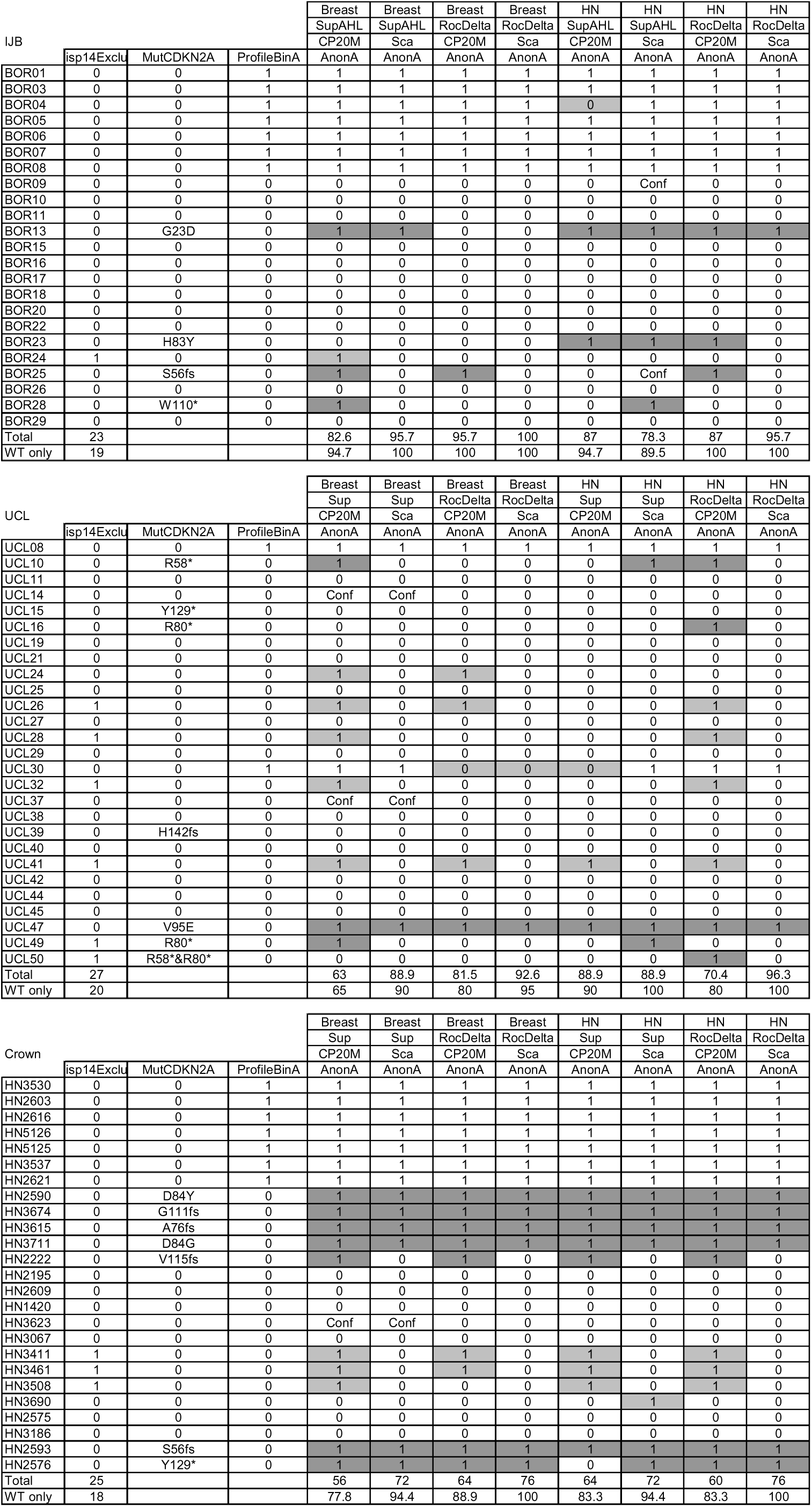
Comparison of the CDK4 profiles of tumors and PDTX models observed with 2D gel electrophoresis or predicted with the tools using gene expression profiles. The tumor or models IDs are reported in the first column. Whether the tumor or the model has an elevated CDKN2A expression exclusively due to the p14-coding mRNA or have a mutated CDKN2A gene is indicated in the second and third column. The observed profiles are reported in the fourth column. The prediction of the CDK4 profiles with the tools described in the 3 first rows are reported in the following columns. The first row on top of the table indicates whether references were built with the breast expression values of the initial study or with the head & neck prototype tumors. The second row on top of the table reports the prediction rule used. With the tool noted Sup, a sample is predicted to have profile A lacking CDK4 phosphorylation if the Spearman correlation coefficient of the comparison between the gene expression values for the 11 genes included in the prediction tool to the reference built and the corresponding average of their expression in reference tumors with A profile is the largest. With the tool noted RocDelta, a sample is predicted to have profile A lacking CDK4 phosphorylation if the difference between the correlation coefficients of the comparison between the gene expression values for the 11 genes included in the prediction tool to the reference built and the corresponding averages of their expression in reference tumors with A or non A profiles, respectively, is above a threshold defined by a ROC analysis in the analysis of the cohort of head & neck tumors from the IJB. Tumors of the IJB cohort or of the UCLouvain cohort as well as Crown Bioscience models are described in the corresponding panels (noted IJB, UCLouvain and Crown, respectively). The total numbers of tumors or models analyzed with or without *CDKN2A* mutation are reported at the bottom of the second column of each tables. The proportion of correct predictions considering the whole tumor or model cohort or considering only tumors or models without *CDKN2A* gene mutations are indicated below each column describing the predicted profile of the analyzed tumors or models. Frame-shift mutants (indels) are noted with the amino acid affected, its position and the code fs. Mutants wherein a codon is converted to a termination codon are noted with the amino acid affected, its position and the symbol *. When the correlation coefficient of the comparison of the unknown sample gene expression profile to the H, L or nonA reference profiles is the same as the corresponding coefficient of the comparison to the A profile reference, the CDK4 modification profile cannot be determined. These cases are noted as Conf in table S2. These cases are considered as false predictions in the computing of the accuracies in the last two rows of the subtables of table S2.

**Supplementary Table S3.**
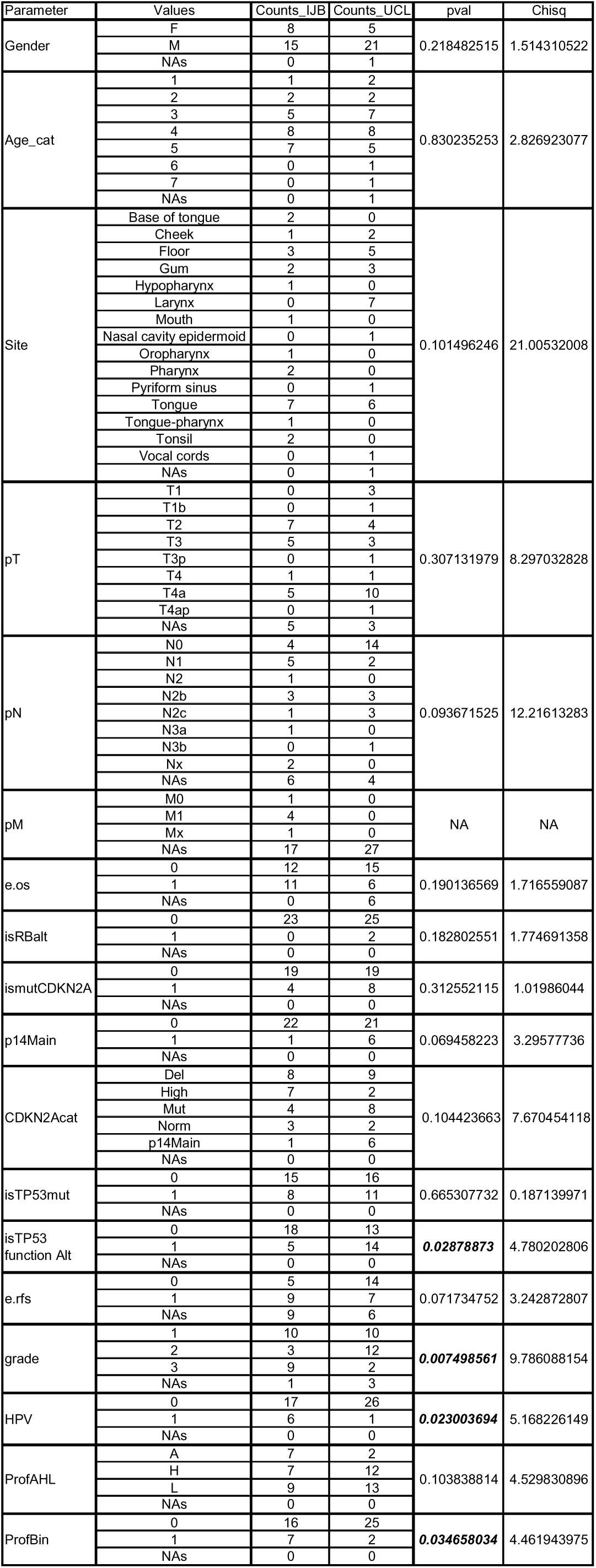
Comparison of the demographic data of the IJB and UCLouvain HNSCC tumor cohorts. The counts of tumor samples with the indicated values of the described clinical features in the final selection of tumors from the IJB and UCLouvain cohorts are indicated in the “Counts_IJB” and Counts_UCLouvain columns, respectively. The NA values correspond to missing data. The Chi-square statistic and the p-value to reject the null hypothesis of independence are reported for the comparison of the proportions of cases in the two cohorts with the observed values of the clinical features considered. The two cohorts differs in all clinical features linked to the HPV status (grade, relapse events, CDK4 profile, *TP53* gene alteration status) due to the lower number of HPV-positive cases in the UCLouvain cohort.

**Supplementary Table S4.** Antibodies used in the study The names, references, sources, characteristics and use of the antibodies included in this work are reported. The clone ID of monoclonal antibodies are reported when indicated.

| Application | Antigen [conjugate] | Provider | Clone | Reference | Dilution |
| --- | --- | --- | --- | --- | --- |
| Immunofluorescence | BrdU | BD Bioscience | B44 | 347580 | 1/50 |
| Immunofluorescence | Mouse IgG [biotinylated] | GE Healthcare | - | RPN1001V | 1/50 |
| Immunofluorescence | Streptavidin [Alexa Fluor 488] | Jackson ImmunoResearch | - | 3016-480-084 | 1/3600 |
| Two-dimensional gel electrophoresis | CDK4 | Santa Cruz Biotechnology | H-22 | sc-601 | 1/200 |
| Two-dimensional gel electrophoresis | CDK4 | Cell Signaling Technology | D9G3E | 12790 | 1/1000 |
| Western blot | Mouse IgG [HRP] | Cell Signaling Technology | - | 7076 | 1/2000 |
| Western blot | Rabbit IgG [HRP] | Cell Signaling Technology | - | 7074 | 1/2000 |
| Western blot | Cyclin A2 | Gift from Tim Hunt | E43.2 | - | 1/20 |
| Western blot | Cyclin D1 | Neomarkers | DCS-6 | MS-210-P | 1/200 |
| Western blot | Cyclin D1 | Invitrogen | DCS-6 | AHF0082 | 1/200 |
| Western blot | Cyclin D3 | Neomarkers | DCS-22 | MS-215-P | 1/200 |
| Western blot | Cyclin E | Thermo Fischer Scientific | HE12 | MA5-14336 | 1/200 |
| Western blot | CDK4 | Cell Signaling Technology | D9G3E | 12790 | 1/1000 |
| Western blot | P-CDK4 | in house production | NB8-AD9 | - | 1/1000 |
| Western blot | CDK6 | Neomarkers | DCS-83 | MS-398 | 1/1000 |
| Western blot | E2F1 | Cell Signaling Technology | - | 3742 | 1/750 |
| Western blot | p14 | Santa Cruz Biotechnology | DCS-241 | sc-53640 | 1/200 |
| Western blot | p16 | Santa Cruz Biotechnology | DCS-50 | sc-65476 | 1/200 |
| Western blot | p21 | Cell Signaling Technology | 12D1 | 2947 | 1/1000 |
| Western blot | p27 | Cell Signaling Technology | D69C12 | 3686 | 1/800 |
| Western blot | pRb | BD Pharmingen | G3-245 | 554136 | 1/500 |
| Western blot | phospho-pRb (S807/811) | Cell Signaling Technology | D20B12 | 8516 | 1/1000 |
| Immunoprecipitation | CDK4 | Santa Cruz Biotechnology | H-22 | sc-601 | 3 µg/reaction |
| Immunoprecipitation | cyclin D3 | Neomarkers | DCS-28 | MS-216 | 3 µg/reaction |
| Immunoprecipitation | cyclin D1 | Abcam | EPR2241 | ab134175 | 3 µg/reaction |
| Immunoprecipitation | p21 | Cell Signaling Technology | 12D1 | 2947 | 3 µg/reaction |
| Immunoprecipitation | CDK6 | Santa Cruz Biotechnology | C-21 | sc-177 | 3 µg/reaction |
| Immunohistochemistry | KI67 | Roche Diagnostics | 30.9 | 790-4286 | - |
| Immunohistochemistry | p16 | Roche Diagnostics | E6H4 | 805-4713 | - |
| Immunohistochemistry (Crown Bioscience PDTX) | KI67 | Abcam | SP6 | ab16667 | - |
| Immunohistochemistry (Crown Bioscience PDTX) | p16 | Cell Signaling Technology | BC42 | 684105 | - |

## Detailed Methods

### Protein analysis and quality criteria of 2D gel detection of CDK4

Chemiluminescence images of the samples were acquired with a VilberLLourmat Solo7S camera and quantified using the Bio1D software (Vilber-Lourmat, Marne-la-Vallée, France). The profile of CDK4 separated by 2D-gel electrophoresis has been characterized previously (Bockstaele, Kooken et al., 2006, Raspe et al., 2017). The spot 2 and spot 3 volumes (corresponding to the two main modified forms of CDK4) were quantified from 16Lbit scans of the 2DLgel immunoblots, with unsaturated signals. Samples for which the signal of the most abundant native CDK4 (spot1) did not reach saturation at an exposure time of 16 minutes the latest were considered to lack sensitivity. Samples were also discarded if the signals started to decrease (eventually due to chemiluminescence substrate exhaustion) before spot2 and/or spot3 were correctly detected. After linear correction of the background, the volume ratio (spot3/spot2) was used to define three types of CDK4 modification profiles: a profile A (for absent phosphorylation) was attributed when the spot3/spot2 ratio was below 0.02; a profile L (for low phosphorylation) was attributed when the ratio lied between 0.02 and 1; and a profile H (for high phosphorylation) was given when the ratio was equal or above 1. The first threshold is the one previously used to classify the thyroid tumor profiles (Pita, Raspé et al., 2023).

As CDK4 is not phosphorylated in quiescent cells, samples without phosphorylated CDK4 and lacking Ki67 positive cells were considered as normal tissue and discarded after examination of the stainings by a pathologist. For 2D gel electrophoresis image documentation, the centers of mass of spot 1 and 2 were localized with the QuPath-0.5.0 software using the appropriate exposure TIF image. The distance between spot2 and spot1 were computed for each gel. Each image was linearly scaled using the magick R package in such a way that the final spot1-spot2 distance of each image equals the average of all distances. The coordinates of Spot2 were checked before a 300 x 200 pixels region of interest centered at the spot2 center of mass was extracted. Samples were also excluded when the spots were not correctly focused and appeared as smears or when the distance between the modified forms of CDK4 was not sufficient. At the start of the study, because of the limited sensitivity of the primary antibody ECL detection kit, the minimal protein concentration required to detect the phosphorylated CDK4 was 0.75 µg/µl. As more sensitive detection kits were available later in the study, less concentrated protein samples were used for the re-analysis of some tumors. To adjust the selection to the increased sensitivity of the new primary antibody ECL detection kits while selecting only signals high enough to detect P-CDK4 with good confidence, we included samples in the study if their 2D gel electrophoresis spot1 (native CDK4) saturated after an exposure time of maximum 16 minutes. If this was not the case, a sample was also included if its spot1 reached at least a maximal height of 40000 and if its spot3 reached a maximal height of at least 2000. In addition, the spot2 (yet unknown modification) and spot3 (phosphorylated CDK4) signals should not decrease. The exception is a sample (UCL10) with saturated spot1 with a decreasing signal only at the last exposure time with a spot3 signal reaching a maximal height of 5000 with the same profile at all exposure times.

### Tumor sample RNA extraction quality criteriae and analysis of RNASeq data

Based on the recommendation of the cDNA library preparation kit, a library was prepared if the minimal concentration of the RNA samples was 10 ng/µl and if the RIN value estimated by a Bioanalyzer analysis reached 5. A sample was included in the study if the proportion of unmapped too short reads did not exceed 35%. Samples were also excluded from the study when the Ki67 IHC failed to reveal tumor cells and when the cumulative relative rank of the expression of key genes with increased expression in dividing cells (*SPDL1*, *ESCO2*, *KIF20A*, *KIF2C*, *MYBL2*, *NCAPH* and *RRM2*) was below 10. These samples were not considered because they mainly included non-proliferative normal tissue wherein CDK4 is not phosphorylated. The pileup function of the R package Rsamtools was used to extract the coverage values at each position of the *CDKN2A* locus from each sample BAM file. Genomic coordinates of the whole locus and all introns and exons coding for p14 and p16 were extracted from the gtf file used for the aligning the reads on the human genome reference. A linear regression between the coverage at each intronic position and their gene coordinates was used to correct the coverage at each exon positions for background. When background estimates were higher than the observed coverage, the value was set to zero. As the best fit between exon 2 coverage and the sum of the exons 1α and 1β coverage was obtained using maximum coverage values, this quantification was prefered. To be able to compare *CDKN2A* gene and exon 1α expression levels in each sample, exon 1α expression was calculated as a fraction of *CDKN2A* gene expression. This fraction was defined by the ratio of maximum coverage values between exon 1α and the sum of exons 1α and 1β maximum coverage values in each of the samples.

To systematically search for mutations in the RNASeq data, a custom pipeline was set up in R. The coordinates of the gene exons was first extracted from the gtf file used for the alignment and completed with the sequence extracted from the corresponding splited fasta file to create a feature reference file. Genes present in the Cosmic Cancer Gene Census were selected and completed by a custom list of other genes of interest. The pileup function of the R package Rsamtools was used to extract the proportions of each nucleotides recorded at each positions defined by the feature reference file. If the coverage at these positions is higher or equal to 10 and if the proportion of any nucleotide different from the nucleotide present in the consensus sequence of the gene of interest at this position is greater or equal to 10%, the proportion of nucleotide (or the indel status), the nucleotide of the consensus gene sequence at the position and the position are recorded. These data are annotated with the amino acid affected and its position in the protein, the type of mutation and the functional consequence of the mutation, if known, using the predictCoding function of the VariantAnnotation library using the BSgenome.Hsapiens.UCSC.hg38 and TxDb.Hsapiens.UCSC.hg38.knownGene data libraries. Synonymous mutations are discarded. Information on functional impact of the protein sequence variations recorded is completed using the Cosmic database and manually completed for genes of particular interest.

### Prediction of the CDK4 modification profile

As previously detailed and characterized (Raspe et al., 2017), a centroid method was used to predict the CDK4 profile whereby the expression profile of 11 genes (including *CDKN2A*) of an unknown sample was compared to three references built by computing, for each 11 genes, the average of their expression among prototype tumors with A, H or L CDK4 profiles. The predicted profile was the one corresponding to the A (no phosphorylated CDK4), H (phosphorylated CDK4 is the main modified form of the enzyme) or L (phosphorylated CDK4 is the minor modified form of the enzyme) centroid with the highest correlation coefficient. For binary profile prediction, references were built with gene expression profiles of prototype tumors with CDK4 profile A or non-A (H or L profiles). As they were initially profiled with the Affymetrix platform (Raspe et al., 2017), the breast tumor references used to predict the CDK4 modification profile were first adapted by using the RNA-Seq expression data acquired with RNA extracted from the same samples. Raw HNSCC tumor RNA-Seq CP20M expression values were compared to these adapted references by Spearman correlation. For the *CDKN2A* gene, these values were also corrected, as detailed above, by the contribution of the p16-specific exon 1α to the expression of the whole *CDKN2A* locus expression. In this case, all samples were scaled by the same factor such as the average expression *CDKN2A* exon 1α of profile A HNSCC tumors was equal to the *CDKN2A* value of the reference for profile A breast or HNSCC tumors. Samples in which native CDK4 was not saturated after an exposure of 16 minutes were excluded as quality and yield of RNA extraction were often low in such samples.

### Immunohistochemistry and tissue section analysis

Immunohistochemical stainings of hematoxylin/eosin (HE), KI67 and p16 were performed at the anatomo-pathology department of the IJB with FFPE tissue sections on a fully automated BenchMark Ultra IHC system (Ventana, Roche Diagnostics, Basel, Switzerland) with the UltraView Universal DAB Detection Kit (Roche Diagnostics), using standard routine protocols using three consecutive 5 µm sections cut from each tumor sample. Pictures were acquired with a NanoZoomer digital scanner (Hamamatsu Photonics, Hamamatsu, Japan) at 40× magnification with the 3 layers option. HE and immunohistochemical (Ki67 and p16) of FFPE tissue from head & neck PDTX model samples were performed at Crown Bioscience using their routine procedure with a Bond RX automatic IHC/ISH system (Leica). The Abcam anti-Ki67 (ab16667) and the Cell Signaling Technology anti-p16^INK4^ (684105) antibodies were used for primary detection revealed using the Bond Polymer Refine Detection kit (DS4800, Leica). Pictures of stained slides were acquired with a Pannoramic Scan Digital Scanner (3DHistech) at 40X magnification.

To illustrate tumors or PDTX models, representative fields of interest (1400X1400 or 140X140 µm) were selected with the Qpath software on the Ki67 staining images. To this end a project was created in Qpath including the Ki67 and the p16 or the Hematoxylin/Eosin staining image of each tumor or model. The 1400X1400 µm and the 140X140 µm region included in the 1400X1400 µm region were first selected in the Ki67 staining image of the project. The centroid coordinates of these regions were recorded. Next, either the p16 or the Hematoxilin/Eosin staining corresponding images were aligned on the Ki67 staining image with the Interactive image alignment function in Qpath. After alignment, the p16 or Hematoxylin/Eosin staining images of the selected aligned regions of interest were exported to ImageJ with the Send region to ImageJ extension of Qpath and saved as tif files. The Affine transform matrix generated by the Qpath interactive image alignment function was recorded. The p16 or Hematoxilin/Eosin staining image was opened in Qpath. Regions of interest were created on this image using the previously recorded coordinates of the corresponding regions in the Ki67 staining image. The Affine transform matrix is applied on these regions of interest with a custom script by the Automate-Script editor function of Qpath. The transformed regions of interest are then aligned on the Qpath frame using the View-Rotate image function of QPath. The rotation angle is recorded. The position of the transformed regions of interest is then manually adjusted to fit with the recorded images obtained before during the alignment procedure. The centroids of these transformed and adjusted regions of interest are recorded. The recorded centroid positions and rotation angles for each tumor or models are inserted using R in a script module master text to create a region of interest export script. This script is applied in Qpath on projects with all Ki67, p16 or Hematoxylin/Eosin staining images with the Automate-Script editor function of Qpath to extract images the regions of interest from the Ki67, p16 or Hematoxylin/Eosin staining images, respectively. These region of interest images are saved as tif files. A scale bar is then added to these images with custom scripts in ImageJ. These resulting images are finally combined with 2D gel electrophoresis images and tumor or model characteristics in R using custom scripts utilizing the Magick R package. This semi-automatic procedure ensure reproducible and totally documented handling of the images.

## Readme File description

The work includes the following files (with their description)

Article_Raspe_et_al_HNSCC_20251104.docx: main article file

Figures_Horizontal_Raspe_et_al_HNSCC_20251104.pdf : all figure in landscape orientation (cover figure, figure 2, figure 3, figure 4, figure EV2 and figure EV3)

Figures_vertical_Raspe_et_al_HNSCC_20251104.pdf : all figure in portrait orientation (Figure 1, Figure EV1 1 to 4, figure EV4 and figure EV5 1 to 5)

Dataset_EV1_Clin_data_20251020.xls: supplementary tables EV1 (clinical and molecular data of tumors)

Dataset_EV2_PDTX_20251020.xls: PDTX selection and molecular data

Dataset_EV3_lines_20251016.xls: cell lines selection and molecular data Dataset_EV4_20251022.xls: HNSCC selection

